# An overview of the National COVID-19 Chest Imaging Database: data quality and cohort analysis

**DOI:** 10.1101/2021.03.02.21252444

**Authors:** Dominic Cushnan, Oscar Bennett, Rosalind Berka, Ottavia Bertolli, Ashwin Chopra, Samie Dorgham, Alberto Favaro, Tara Ganepola, Mark Halling-Brown, Gergely Imreh, Joseph Jacob, Emily Jefferson, François Lemarchand, Daniel Schofield, Jeremy C Wyatt, NCCID Collaborative

## Abstract

The National COVID-19 Chest Imaging Database (NCCID) is a centralised database containing chest X-rays, chest Computed Tomography (CT) scans and cardiac Magnetic Resonance Images (MRI) from patients across the UK, jointly established by NHSX, the British Society of Thoracic Imaging (BSTI), Royal Surrey NHS Foundation Trust (RSNFT) and Faculty. The objective of the initiative is to support a better understanding of the coronavirus SARS-CoV-2 disease (COVID-19) and development of machine learning (ML) technologies that will improve care for patients hospitalised with a severe COVID-19 infection. The NCCID is now accumulating data from 20 NHS Trusts and Health Boards across England and Wales, with a total contribution of approximately 25,000 imaging studies in the training set (at time of writing) and is actively being used as a research tool by several organisations. This paper introduces the training dataset, including a snapshot analysis performed by NHSX covering: the completeness of clinical data, the availability of image data for the various use-cases (diagnosis, prognosis and longitudinal risk) and potential model confounders within the imaging data. The aim is to inform both existing and potential data users of the NCCID’s suitability for developing diagnostic/prognostic models. In addition, a cohort analysis was performed to measure the representativeness of the NCCID to the wider COVID-19 affected population. Three major aspects were included: geographic, demographic and temporal coverage, revealing good alignment in some categories, e.g., sex and identifying areas for improvements to data collection methods, particularly with respect to geographic coverage. All analyses and discussions are focused on the implications for building ML tools that will generalise well to the clinical use cases.

## Background & Summary

Radiology has played a significant role during the pandemic, informing our understanding of the COVID-19 disease (Hosseiny 2020, Kooraki 2020, Shi 2020, Lee 2020) and guiding decision making along care pathways. Clinicians have identified characteristic features of COVID-19 acute respiratory distress from thoracic imaging studies; such features can be used to differentiate COVID-19 patients from those suffering other respiratory conditions (Shi 2020, Chung 2020, Kanne 2020). However, these differences in disease manifestation are often subtle (Cleverley 2020) and may be more quantitatively delineated using computational methods.

One corollary of the widespread adoption of radiology during the pandemic is the accumulation of large volumes of clinical imaging data spread across hospital sites throughout the UK. The National COVID-19 Chest Imaging Database (NCCID) was established to collate this mass of X-ray, CT and MRI scans into an accessible imaging database. The end goal of the NCCID is to facilitate researchers and technology developers in the creation of fair, effective and generalisable ML technologies that can support diagnosis, prognosis and risk stratification of the COVID-affected population, ultimately aiding clinicians to improve patient outcomes.

The initiative was formed as part of the NHS AI lab’s mission of enabling the safe adoption of AI technologies in the NHS (NHSX, AI lab n.d.) and was successfully set up through partnerships with the Royal Surrey NHS Foundation Trust, the British Society of Thoracic Imaging (BSTI) and Faculty, an AI technology company. This combination of data processing and clinical expertise has been leveraged to create a data warehouse comprising pseudonymised thoracic imaging and relevant clinical data points for thousands of patients across the UK.

The legal basis for the NCCID is provided by the notice under regulation 3(4) of the UK National Health Service (Control of Patient Information) Regulations 2002 (COPI Notice), and ethical approval was obtained for the NCCID to operate as a research database by the UK Health Research Authority. Further information on the NCCID’s remit and rationale are described in an article in the European Respiratory Journal (Jacob 2020).

A portion of the data is transferred to the training set, which contained 24,465 imaging studies from 7,685 patients at time of writing (latest figures can be found on the [NCCID information page](https://nhsx.github.io/covid-chest-imaging-database/stats.html)). This training data is available to any users, including software vendors, academics and clinicians, via a rigorous Data Access Request (DAR) process. Applications are adjudicated by an independent committee based on several factors including but not limited to relevance to COVID-19 and compliance with information governance regulations. The required paperwork and additional instructions are detailed [on the website](https://nhsx.github.io/covid-chest-imaging-database/index.html#how-to-request-access-to-data).

The remaining portion of data is allocated to the validation set, which is protected as a hold-out set for NHSX to conduct future performance assessments of COVID-19 chest-imaging AI technologies, ensuring that they are safe and effective before testing in a real-world clinical setting. Results presented in this paper are solely focused on the training data, in order to maintain the integrity of the validation data as a hold-out benchmarking tool.

This article aims to describe key characteristics of the data and indicate its usefulness for developing algorithms that can support COVID-19 diagnosis and prognosis from chest images. The work was conducted on pseudonymised data within the existing NHSE AWS cloud infrastructure for the NCCID. To preserve the privacy of individuals, suppression of small numbers has been implemented throughout the paper. Suppressed data is indicated within plots and tables by the presence of an asterisk (*) for categories containing less than 7 individuals.

As the data is submitted in two parts - the images themselves, and the clinical data separately the analysis has naturally been structured in this manner with an additional investigation of how the geographic, demographic and temporal coverage of the dataset compares with publicly available datasets for the wider COVID-affected population. The implications of these findings for developing algorithms related to COVID-19 is provided in the Discussion, alongside a list of future aims that have been identified to improve the dataset.

## Methodology

### Database setup

Figure 1 provides an overview of the data collection pipeline for the NCCID warehouse, which can be broadly broken down into the following stages:

**Figure 1.**
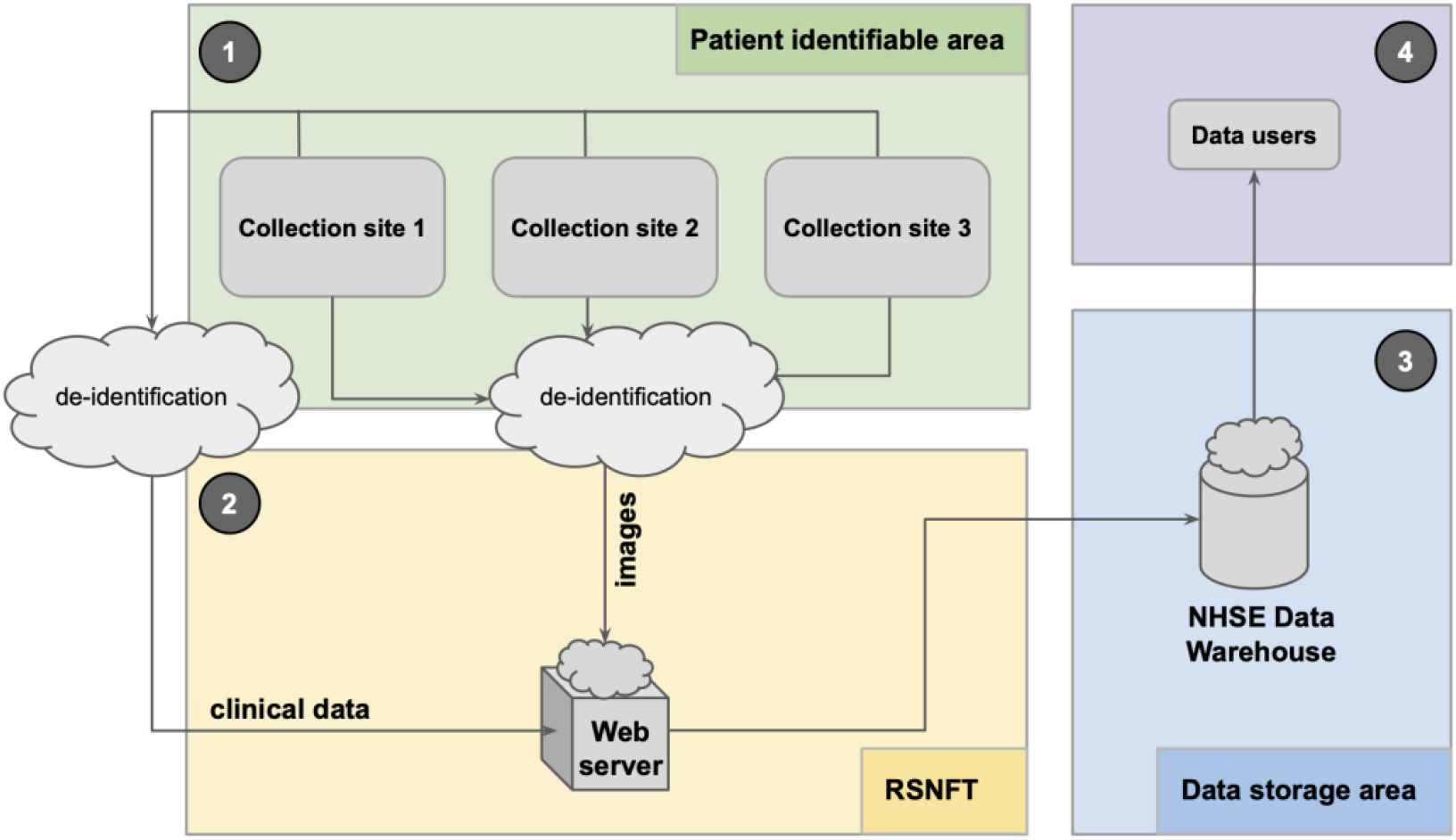
diagram of the data collection pipeline for the NCCID warehouse.

1. NCCID participating collection sites (hospitals) are requested to contribute imaging data for patients that have undergone a real-time Reverse Transcription Polymerase Chain Reaction (RT-PCR) test for COVID-19. In addition to the images, two spreadsheets with different fields for the positive and negative cases are populated to capture accompanying clinical data (see *clinical data* and *supplementary resources* for more information).
2. The Scientific Computing Team at RSNFT have established a dedicated node on Sectra’s Image Exchange Portal (IEP) for receiving the images. IEP is a widely used network for sharing images between hospitals. The images are received by a SMART (Secure Medical-Image Anonymiser Receiver for Trials) box in Random Access Memory (RAM) and de-identified before writing to disk, ensuring that no patient identifiable information leaves the sites. The clinical data spreadsheet is also de-identified by means of a common pseudonym, generated via a one-way hashing algorithm combined with a complex salt and uploaded to a web portal. Upon receiving images and clinical information, RSNFT links the two sources using the pseudonym. Patient’s unique digital identifiers (NHS number or equivalent for devolved nations) are also encrypted using an Advanced Encryption Standard (AES) algorithm and a complex salt to allow linkage with other national-level datasets.
3. The data is transferred to a central NCCID data warehouse hosted inside NHS England’s (NHSE) Amazon Web Services (AWS) infrastructure, designed and implemented by Faculty and NHXS. The warehouse is backed by a single Simple Storage Service (S3) bucket within a separate sub-account under NHSE’s AWS organisation. All data within the S3 bucket is encrypted at rest using AES-256 encryption. Data is regularly split into training and validation sets based on a randomisation of patients: once a patient has entered the training or validation set, any new images for that patient are automatically added to the same set. The codebase for warehouse infrastructure is open-source (see *supplementary resources*).
4. Data users that have been approved through the DAR process can access the training set. Image files are available in DICOM format, and clinical data is stored as JSON files. AWS credentials for the S3 bucket are provided to an organisation via an encrypted communication. Further support, including guidelines and code for access the data are provided through the [information site](https://nhsx.github.io/covid-chest-imaging-database/data_access.html).

#### Inclusion criteria

The inclusion criteria for individuals within the NCCID database are as follows:

- The person has undergone a COVID-19 swab test (RT-PCR). The outcome of the test may have been positive or negative. Some individuals may have undergone multiple swab tests;
- The person has undergone chest imaging in the three weeks before or after the swab.

The positive cohort consists of the individuals that returned one or more positive swab tests. All imaging data associated with a positive patient’s COVID-19 hospital episode have been requested. To provide insight on longitudinal risk factors, historical images up to January 2017 are also requested.

The negative cohort consists of individuals for whom all acquired swab tests return negative. This may differ from some clinical databases where the control cohort represents healthy individuals but was deemed the correct method for curating a dataset that could train the most useful diagnostic models that differentiate COVID-19 characteristic features from other respiratory conditions. Thoracic images acquired within the six-week window surrounding the test are requested.

Although the status of a patient’s RT-PCR swab test serves as a proxy for ground truth, users should be aware of the limitations of these labels. In particular, this method of testing has a relatively low sensitivity score, where estimates range from *0*.*71-0*.*98* (Watson 2020), this causes the false omission rate to be quite high. In addition, the probability of having a COVID-19 infection is higher in those attending hospital with respiratory symptoms, than for the general public. Given these factors, data users should expect the negative cohort to contain a non-negligible portion of mislabelled positive patients. Additional clinical assessment of the images may be required to improve the accuracy of labels.

### Imaging data

The NCCID is a continually growing asset, as such, all subsequent figures and analyses reported in this paper refer to the training data as of 29 October 2020 (unless otherwise stated). On this date, the NCCID training dataset contained data for 7,500 patients; Table 1.1 details how this cohort is split by control/disease and data availability. There were 1,307 patients with clinical data only due to the fact that the accompanying images had not yet been uploaded by the PACS teams.

**Table 1.1.**
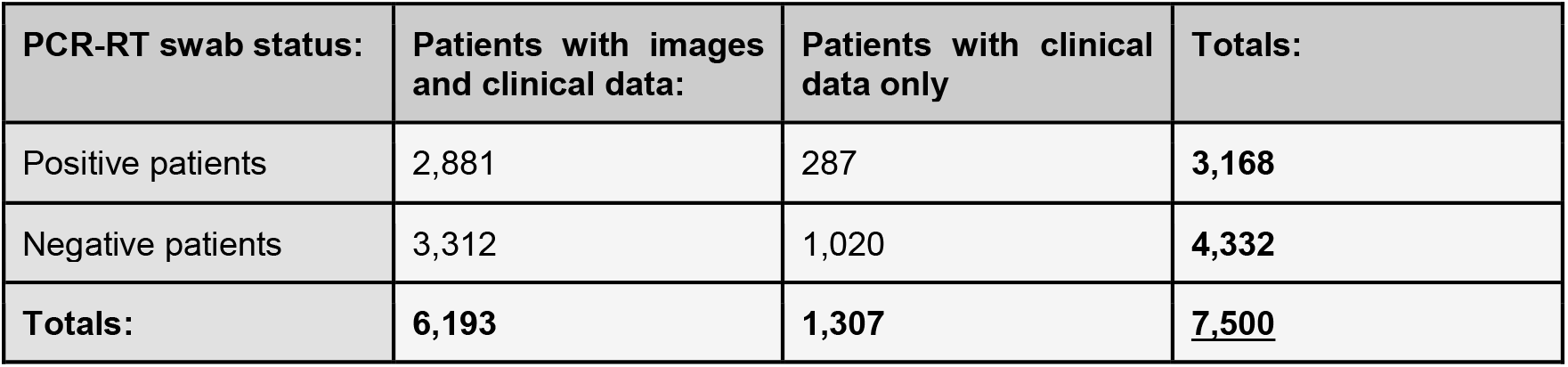
Breakdown of patient cohorts

Table 1.2 details the image modality breakdown for the patients that have had their imaging data uploaded to the training dataset. The majority of the image studies (see glossary in Appendix A for definition) in the NCCID are X-rays, followed by CTs. Only a small number of MRIs, *17*, have been submitted, therefore MRI data is excluded from further analysis. A single patient may have multiple studies within the NCCID, for instance, if multiple diagnostic scans were taken during their treatment pathway or historic scans were provided (see *image characteristics* section for more details).

**Table 1.2.** Modality breakdown of image studies by patient cohort

| PCR-RT swab status: | No. of X-ray studies | No. of CT studies | Totals |
| --- | --- | --- | --- |
| Positive patients | 11,725 | 1,565 | 13,294 |
| Negative patients | 5,532 | 1,112 | 6,651 |
| **Totals** | **17,257** | **2,677** | **19,945** |

### Clinical data

The NCCID sites have been asked to provide additional clinical information alongside imaging data for any patients that have tested positively for COVID-19 via the RT-PCR swab test. The intended purpose of this additional information is to provide researchers with insight into potential causal risk factors, such as comorbidities, as well as potential variables that indicate severity of disease. The clinical data can be broken down into five broad categories:

1. *Demographic information* - age, sex, ethnicity. This data is discussed in detail in the demographics section.
2. *Important dates* such as swab dates, image dates and date of admission.
3. *Patient medical history*, specifying any pre-existing conditions, and the current use of some drugs such as blood pressure medications.
4. *Admission metrics*, detailing the condition of the patient on admission to hospital i.e., blood pressure, lymphocyte count, partial pressure of O2 etc.
5. *COVID information*, pertaining to how the patient was treated (intubation, admitted to ITU), the results of their RT-PCR-tests, the severity associated with their chest X-ray (BSTI n.d.), and their ultimate COVID and mortality status.

For patients in the control cohort, only a subset of this information was requested: patient pseudonym, submitting centre, date of RT-PCR, and result of RT-PCR. This decision was made to reduce the burden on busy ward staff during the pandemic. Schemas for both spreadsheets are available through the supplementary resources section.

Initial investigation of the clinical data revealed several data quality issues, as can be expected during a pandemic when resources and time are understandably limited. Issues included: non-numeric values, such as blank spaces reported for numeric fields; inconsistency of date/time formats with some entries in US (*month-day-year*) versus UK (*day-month-year*) format; mismatch in format for reporting categorical data (e.g., *M, F* for *Male, Female* versus *0, 1*); different sites using different unit scales to report clinical metrics, e.g., *mg/L* versus *ng/L*. To address many of these issues a data cleaning pipeline was created and made publicly available to data users, alongside additional details on the data quality issues, and guidance on the expected format of the clinical data fields (see supplementary resources section).

Missing values in the demographic data were backfilled using a segmentation dataset provided by NHS England and Improvement (NHSE&I) for ethnicity data (internal resource, citation pending), and DICOM header information for sex and age. Making these sensitive attributes available to users is vital for measuring and facilitating equality of care, particularly through bias mitigation of ML models. As such, the additional source of ethnicity data has also been made available to data users.

The results that are reported in this paper are based on the cleaned data for which known errors, such as non-numerical entries have been removed. Text input has been parsed to extract embedded numeric values, and categorical values have been mapped to standard schemas. Issues arising from ambiguous dates (i.e., *03/04* vs *04/03*) and mixed measurement units have not been fully rectified by the cleaning pipeline and may persist.

## Analysis of training data

The following analyses are provided to aid data users in understanding the suitability of the NCCID training dataset for developing diagnostic and prognostic algorithms based on COVID-19 chest imaging:

1. *Clinical data completeness*: assess the completeness and quality of the clinical data, particularly in relation to pertinent information (e.g., comorbidities, disease severity, outcomes) that can provide additional training variables or labels for ML models.
2. *Imaging characteristics*: considers the availability of historical data for longitudinal studies, the implications of the timing of image acquisition along care pathways, and potential model confounders such as the scanner type.
3. *Cohort analysis*: to inform NCCID users of any potential biases in the training dataset that could impede their ability to develop fair, effective, and generalisable AI models. To achieve this, we compared the geographic, demographic, and temporal distributions of patients in the NCCID with publicly available datasets, measuring how far the data is representative of the wider population that has been affected by COVID-19.

The subsequent sections follow the structure of the above three categories, each containing a description of the methodology (if applicable) alongside the key results. The implications of these findings for building ML models are elaborated in the discussion section.

### 1. Clinical data completeness

To understand the utility and limitations of the clinical data with respect to developing diagnostic or prognostic AI models, we assessed the completeness of each field in the four categories: *important dates, patient medical history, admission metrics*, and *COVID information*. Completeness was quantified in terms of the percentage of null and not-null values submitted for each field across all COVID-positive patients.

Figures 1.1 A-D show the completeness of the clinical data after applying the cleaning pipeline (see the clinical data methodology section). For each field of the clinical data, the percentage of entries with non-null values are shown in orange against the percentage of null values in blue. The data exhibits varying degrees of completeness with several well-reported fields present in over *80%* of patients, but the majority of fields are between *0%-50%* complete. The subsequent subsections investigate each plot more closely.

**Figure 1.1.**
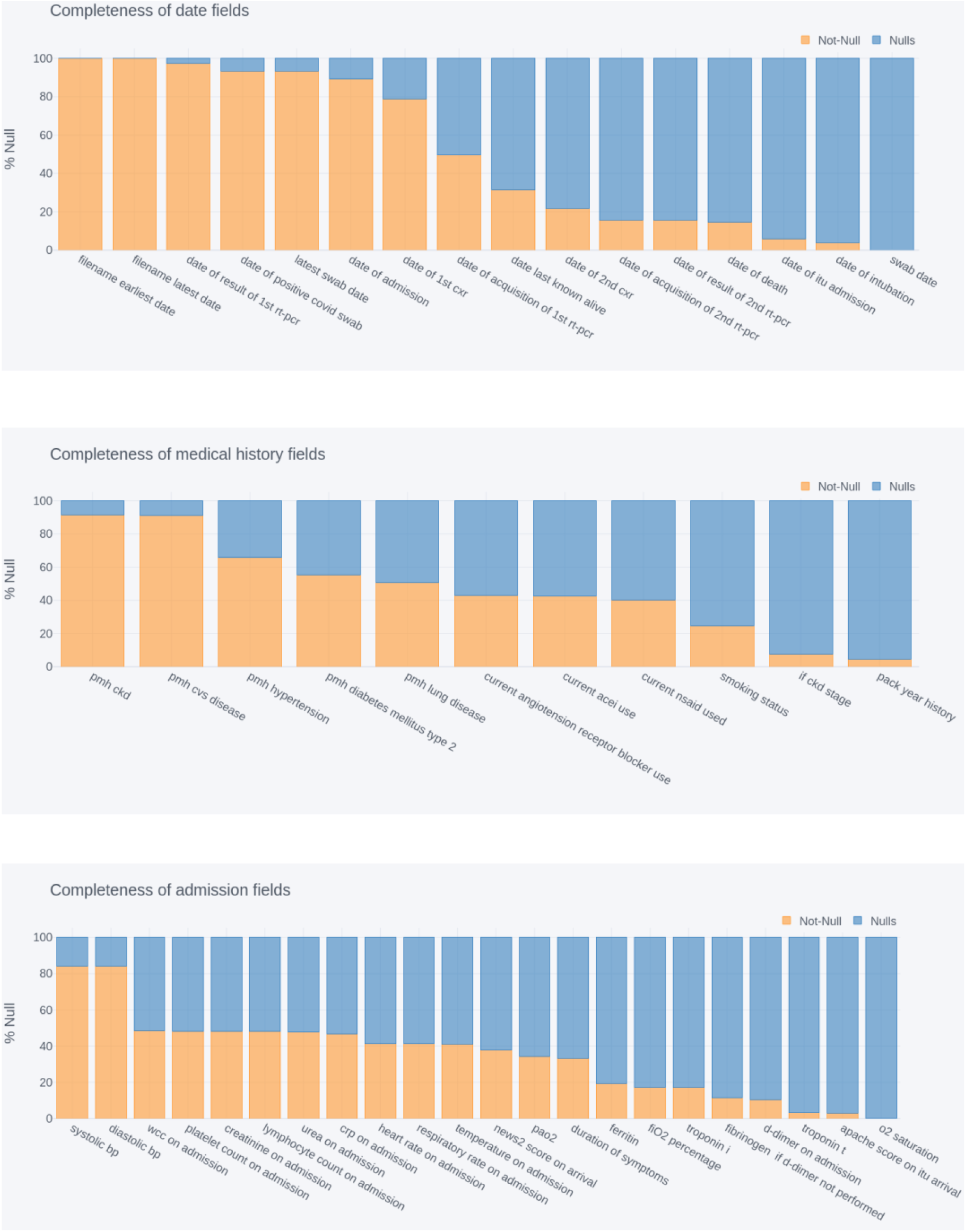
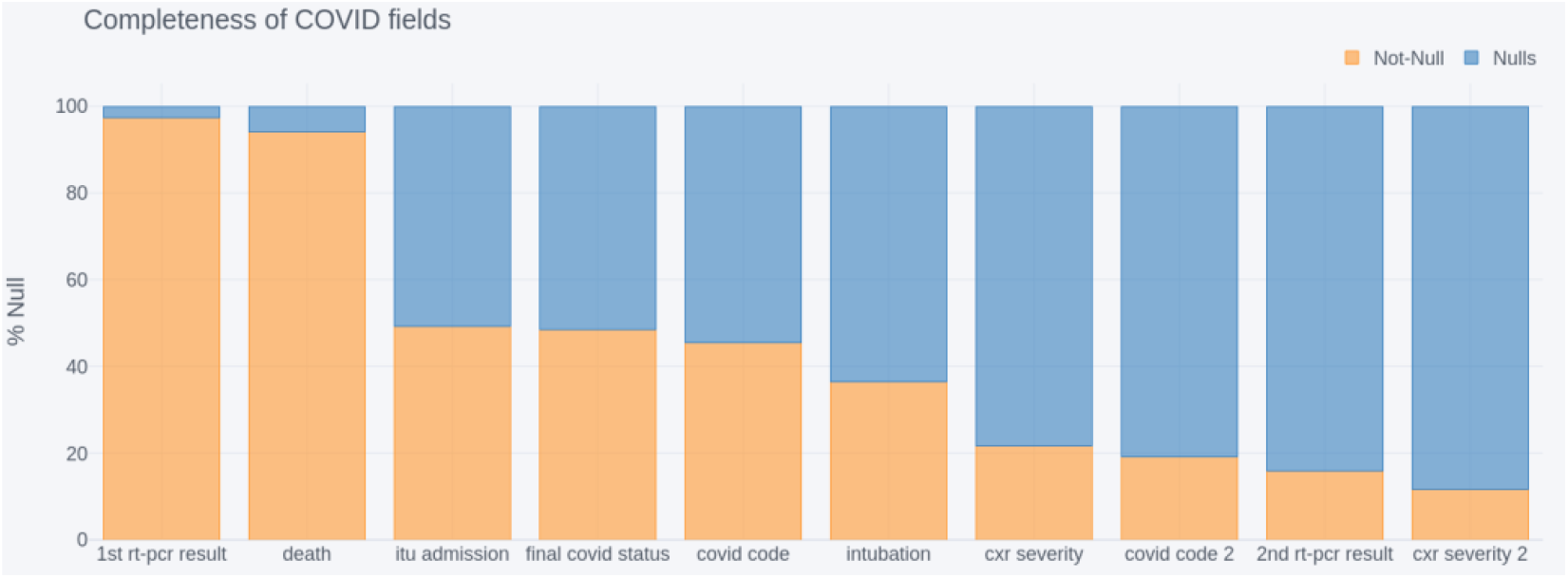
completeness of clinical data fields related to (A) dates, (B) patient medical history, (C) symptoms on admissions and (D) COVID-related information.

#### Dates

The date of *1st PCR result, positive COVID swab, latest COVID swab, admission*, and *1st chest X-ray (CXR)* were well reported, with *79-97%* coverage, whilst dates of *subsequent PCR tests/results, X-rays, ITU admission, intubation* and *death* were present for just *4%-50%* of patients. Coverage for *date of death* increased from *14*.*6%* to *66%* when limiting analysis to the subset of patients for whom the death status had also been reported as positive.

#### Medical history

The presence of *cardiovascular disease* (*CVS*) and *chronic kidney diseases* (*CKD*) were both reported for approximately *90%* of patients. The presence of other pre-existing conditions, *hypertension, type 2 diabetes mellitus*, and *lung diseases* were reported for *66%, 55%* and *51%* of patients, respectively. The use of *angiotensin receptor blockers, ACE inhibitors* (*ACEI*), and *non-steroidal anti-inflammatory drugs* (*NSAID*) were known for between *40-43%* of patients. The patient’s *smoking status* (*never, previous, current*) was known for *25%* of patients, with the *packs per year history* known for *4*.*4%*, increasing to 25% when filtering for patients with *current* or *previous* smoking status. Finally, the stage of chronic kidney disease (*if CKD, stage*) was available for *7*.*5%* of patients overall, rising to *49%* in the subset in which CKD is reported.

For all of these fields other than *pack year history* and *CKD stage*, the reporting includes the negative status of not having the condition. Missing values include that the presence of the condition was marked as *unknown* or left blank.

#### Admission metrics

Of the clinical measurements recorded when a patient is admitted to hospital, blood pressure (systolic and diastolic) was available for *84%* of patients and was by far the most complete field in this category. The majority of remaining fields were reported for between *33-48%* of patients. However, *Ferritin, FiO2, Troponin I, Fibrinogen*, and *D-dimer* were reported for *10-19%* of patients, and *Troponin T, APACHE score* and *O2 saturation* for only *1-3%* of patients.

#### COVID information

The most complete COVID information by far was the *result of the 1st PCR test* and *death status*, which were present for *97%* and *94%* of patients respectively. *Admission to ITU, final COVID status* and *COVID code* were reported for *45-49%* of patients, and *use of intubation* for *36%*. Beyond these the completeness of the fields declined, with *chest X-ray severity* data available for *21%* of patients, *COVID code 2* for *19%, result of second PCR test* for *16%* and *chest X-ray severity 2* for *11%*.

### 2. Image characteristics

This section is designed to inform users on general characteristics of the image data whilst also highlighting potential confounders that might hinder the ability to build effective AI models.

Subsequent sections of the analysis utilise the DICOM header tags associated with image files, these tags were read using open-source package Pydicom (Pydicom n.d.). MRI images are excluded from all analyses due to low numbers in the database at the time of analysis.

#### Historic and acute images

Both acute (related to COVID-19 hospital admission) and historic image studies (up to January 2017) are available for a subset of the NCCID patients. Historic image studies may be used to infer longitudinal risk factors or decouple the effects of pre-existing pathologies from COVID-related symptoms.

Figures 2.1 shows the distributions of the number of historical/acute/total X-ray (A) and CT (B) studies per COVID-positive patient. This number was calculated based on the *date of admission* and the DICOM *StudyDate* (*0008, 0020*), where a study was considered acute if it occurs on or after the admission date and historic otherwise. *Date of admission* was available through the clinical data for *n=2,826* COVID-positive patients; reported results are based on this sample size. In both sets of boxplots, outliers are indicated by dots outside the limit of the plot whiskers and whiskers correspond to Q1 or Q3 +/−1.5*iqr (interquartile range).

**Figure 2.1:**
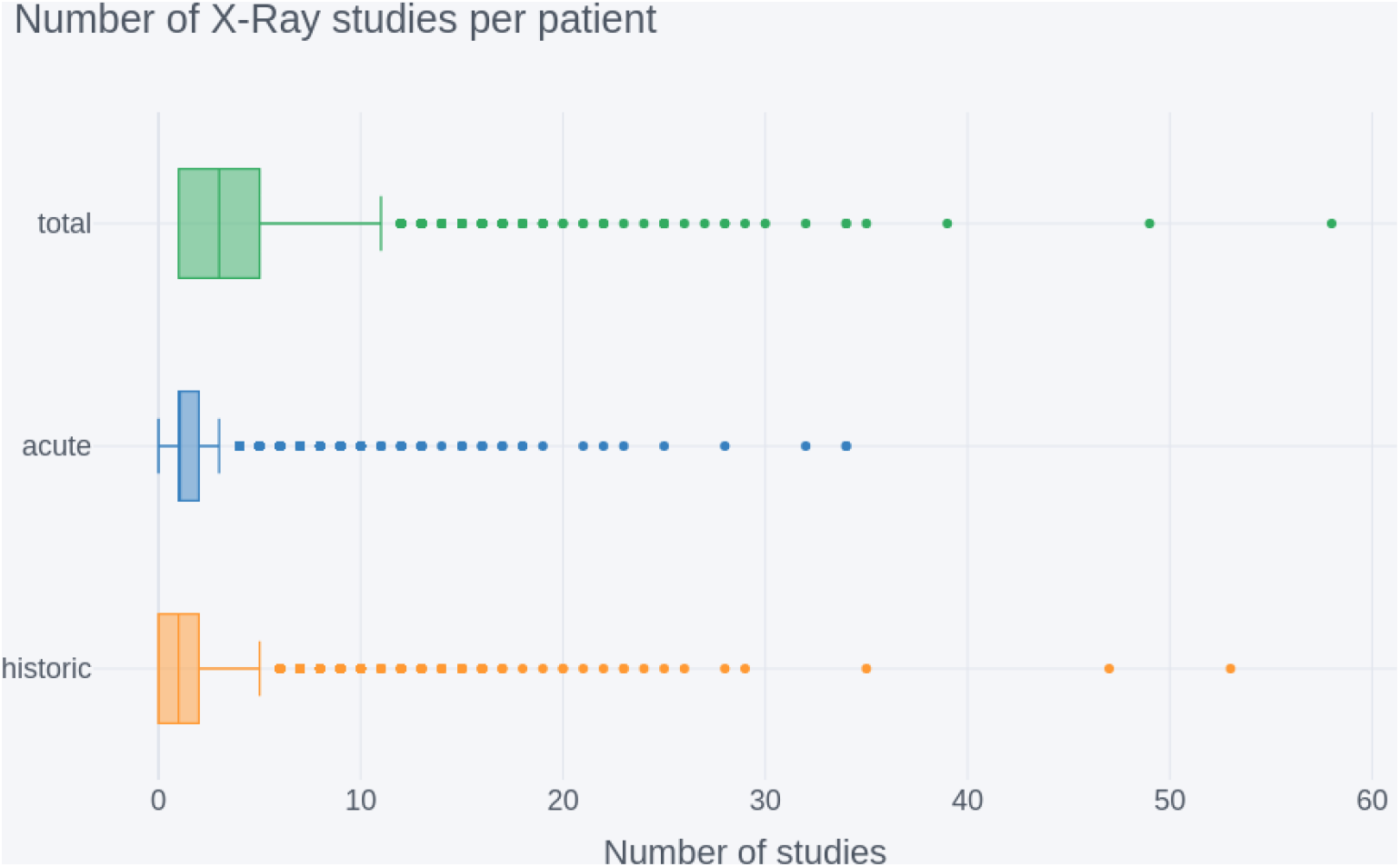
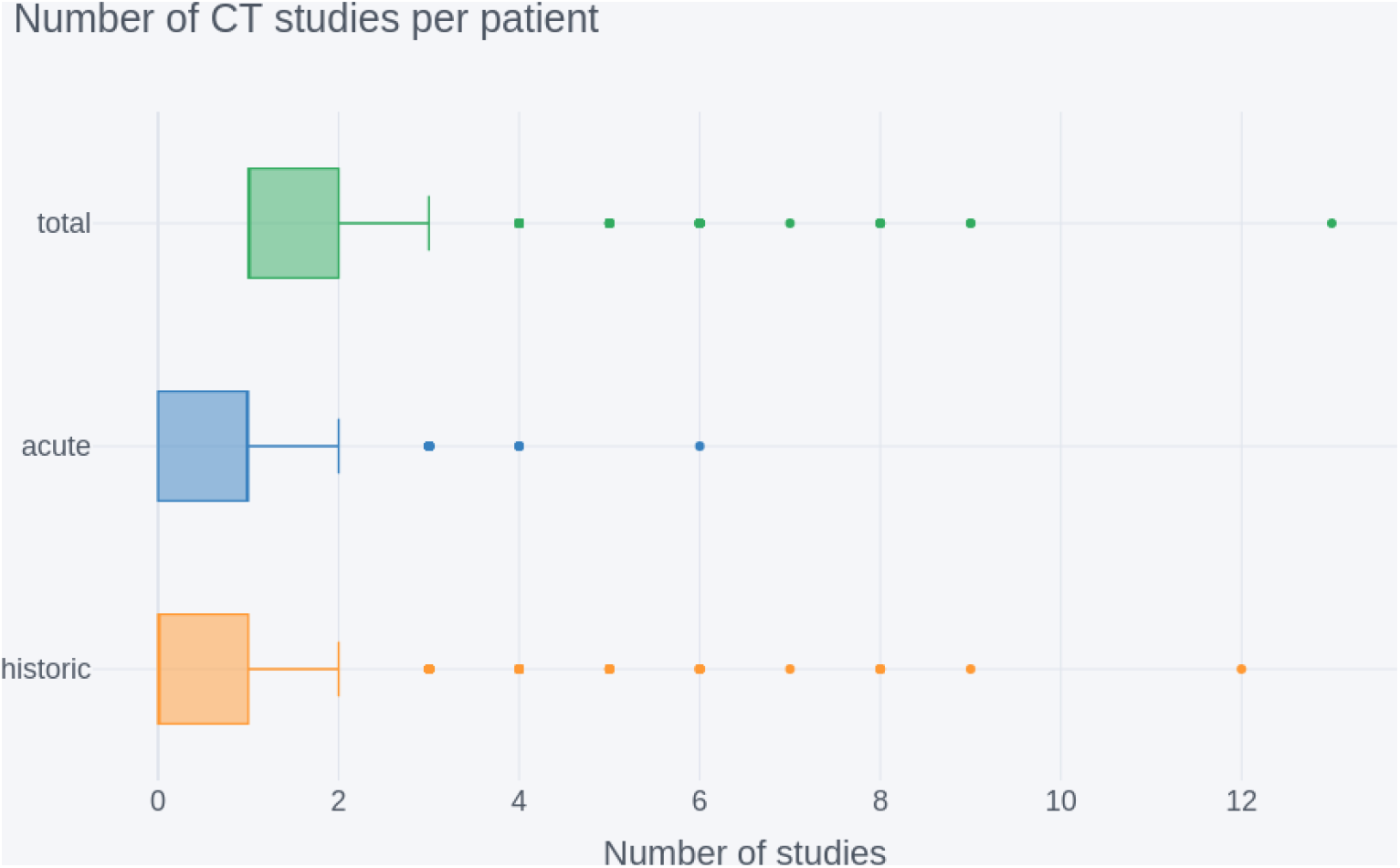
Number of historical/acute/total image studies per NCCID COVID-positive patient (n=2, 826) for (A) X-rays and (B) CTs.

The total number of CTs per patient was *median=1, iqr=1-2*, this was lower than for X-rays (*median=3, iqr=1-5*). This consequently resulted in lower availability of acute CT studies, *median=1, iqr=0-1, max=6*, and even lower availability of historic CT studies, *median=0, iqr=0-1*, but with a handful of patients having *2-12* studies. For X-rays the median number of acute studies per patient was *1*, similar to CT but the *iqr=1-2* is higher, indicating that patients are more likely to have multiple X-rays taken in the acute setting. There was also more historic data available for X-rays, with a *median=1, iqr=0-2*.

#### Acquisition timing

The timing of imaging acquisition along the patient treatment pathway was investigated to understand if different modalities were used for differing purposes in the clinical setting. Two time lags were compared across X-ray studies and CT studies:

1. *days*_*swab to image*_ = *date*_*image*_ - *date*_*positive swab test administered*_
2. *days*_*symptom onset to image*_ = *date*_*image*_ - (*date*_*admission*_ - *days*_*duration of symptom*_)

Image dates were established from the *StudyDate* field of the DICOM headers and lags were calculated based on the first image after the admission date of each patient. This limited analysis to the images taken during the patient’s treatment for COVID-19 in the acute setting. Box plots are used because of the skewed nature of timing data. The distributions of these lags are shown for X-ray (orange) and CT (blue) scans in Figures 2.2 A and B.

**Figure 2.2.**
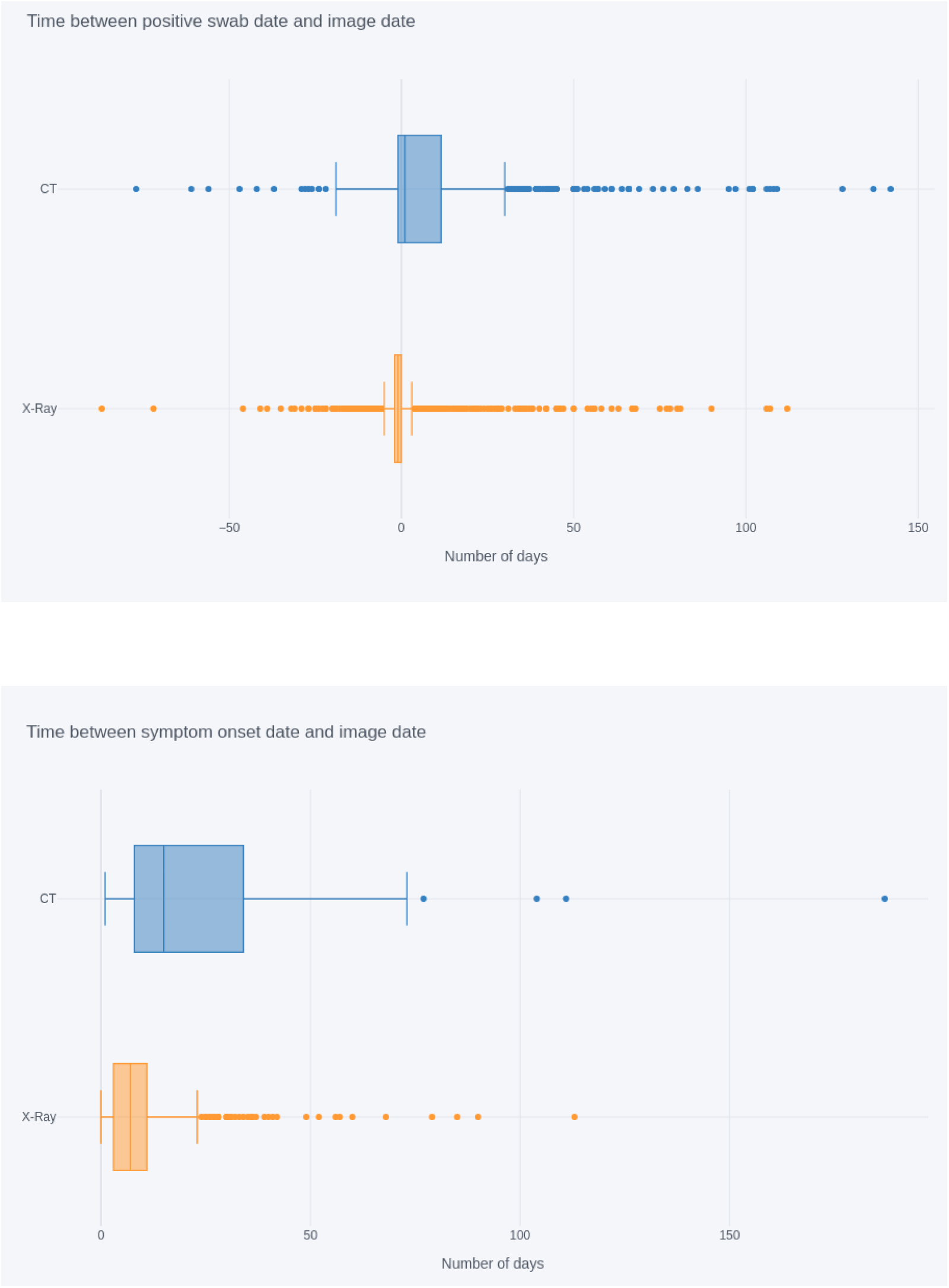
(A) Number of days between the patient’s RT-PCR swab test and the image acquisition (n_XRAY_= 2,410, n_CT_= 507) and (B) Number of days between patient symptom onset and image acquisition (n_XRAY_= 803, n_CT_= 133)

For A), the median offset between swab date and study date was ***-****1* day for X-rays and *+1* day for CT scans. The high number of *-1* day lags for X-ray shows that the majority of X-rays had been taken before a patient’s COVID-19 status was known. The overall distribution across X-rays was far narrower, with an *iqr= −2-0* compared to *iqr= −1-12* for CTs. This suggests that the timing of X-rays is very consistent across patients, whereas longer tails in the CT distribution indicates more variance of usage between patients.

Both modalities display outliers with large negative offsets. These negative offsets suggest that some patients had images taken up to *87* days prior to the positive RT-PCR swab. In practice, the majority of these cases are likely driven by data quality issues surrounding ambiguous dates, such as *03/10* vs *10/03* (see *clinical data* section of methods).

The delay between onset of symptoms and image dates tell a similar story to the above. X-rays had a median offset of *7* days (*iqr = 3-11* days), whilst CTs had a median offset of *15* days and a wider *iqr = 8 - 34* days. Although calculated on a smaller subset of studies (*936* compared to *2917*) for which duration of symptoms data was available, this analysis corroborates the hypothesis that X-rays were consistently used earlier in the care pathway, potentially as diagnostic aids.

#### Scanner types

To investigate the variety of medical imaging equipment within the NCCID database, two analyses were performed:

- Study counts by machine manufacturer were generated using the *Manufacturer* attribute (*0008, 0070*) from the DICOM headers.
- Study counts for model types available within each manufacturer were generated through the combination of DICOM attributes *Manufacturer + Manufacturer’s Model Name* (*0008, 1090*). This combined attribute is hereby referred to as *model*. The results for this additional breakdown are provided in Appendix B.

In both cases, all available DICOM tags were read from each X-ray image file in a study, but only from the first file of each CT study, as the DICOM attributes of interest were the same across all files in a given CT study. Studies for the positive cohort were filtered to exclude historical data based on DICOM *Acquisition Date* (*0008, 0022*) and *date of admission*.

##### Manufacturers

The counts of scanner manufacturers across NCCID positive (orange) and control (blue) cohorts are displayed in Figure 2.3, where ordering of manufacturers is based on the total counts (positive+negative). The total, non-historic, study counts across all manufacturers were *11,086* (*positive=5552, negative=5534*) for X-ray and *1746* (*positive=634, negative=1112*) for CT.

**Figure 2.3.**
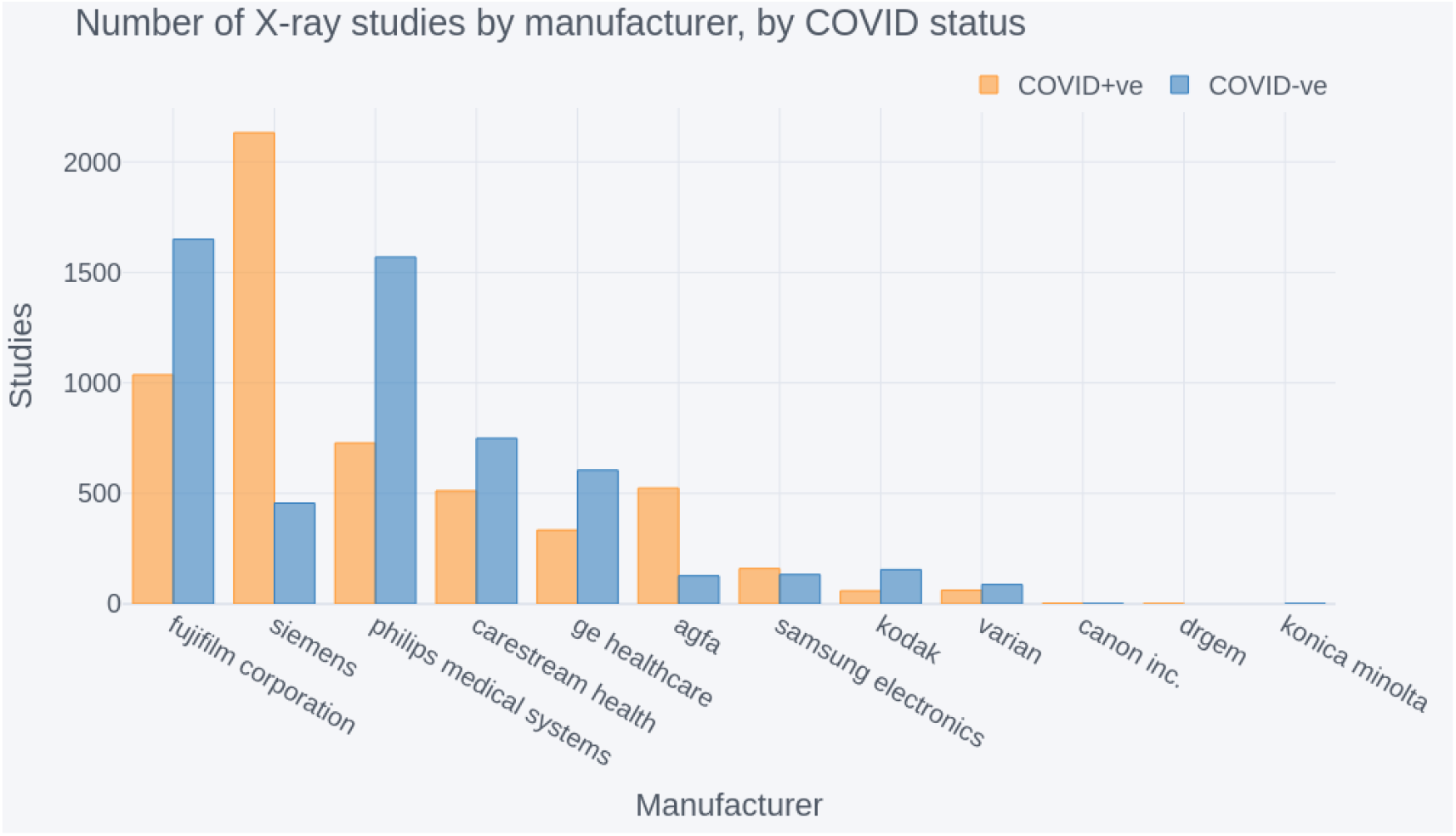
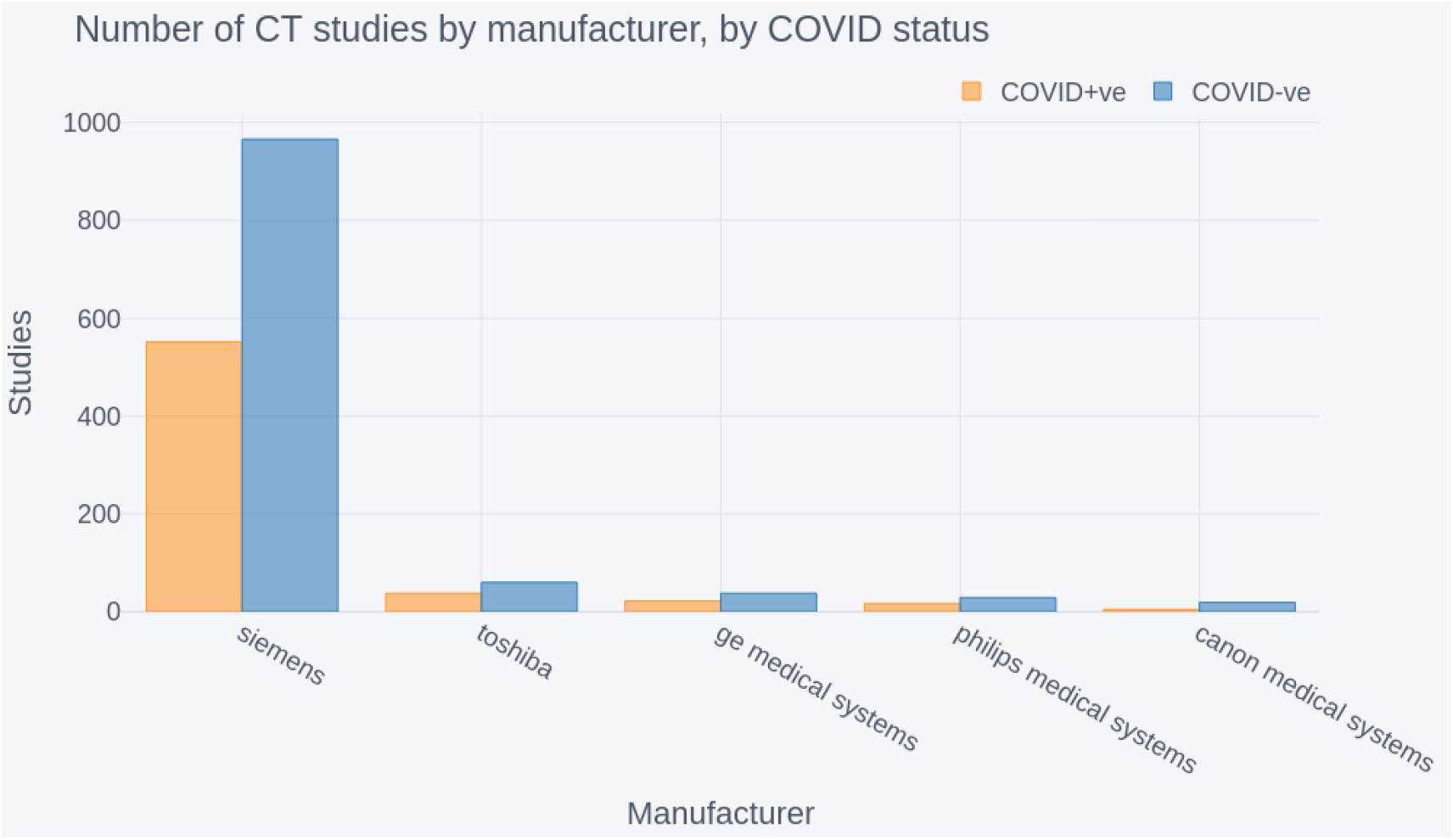
Number of COVID-positive and negative (A) X-ray studies by manufacturer and (B) CT studies by manufacturer. In both cases the manufacturers are ordered by highest to lowest total (positive+negative) number of studies

The largest suppliers for X-rays were *Fujifilm, Siemens* and *Philips Medical Systems*, which contributed *2687, 2588* and *2297* studies each. The next largest supplier was *Carestream Health*, with *1261* studies, after which the number of studies steadily declined for the remaining *8* suppliers. In the case of CT studies, *Siemens* far outweighed the other *4* providers, accounting for *1518* studies.

All X-ray and CT manufacturers had studies for both positive and negative patients. However, some manufacturers, such as Siemens, had significantly more studies in one of the two groups.

##### Portable versus stationary

It was suspected that X-ray data in the NCCID originates from a combination of portable and stationary machines. This was partly a consequence of operational restrictions caused by the pandemic, where portable scanners were easier to regularly disinfect and could be transported to dedicated COVID-19 wards as part of infection control procedures (Kooraki 2020). As such, the use of portable machines was expected to be more prevalent in the COVID-positive cohort of the NCCID.

The percentage of portable scanners was estimated to investigate the presence of potential model confounders caused by e.g., lower image resolution in portable scanners:

- Studies with references to portable, e.g., *CHEST PORTABLE* in the *Body Part Examined* attribute (*0018, 0015*) were counted. Different variations were mapped e.g., *PORT CHEST* to *CHEST PORTABLE*. Studies that did not include any reference to portable in this attribute were assumed to originate from stationary scanners.
- Counts were then adjusted by taking the unique set of eight models from the above step (highlighted in Table B.1) and extrapolating the *portable* status to all studies acquired on these models, under the assumption that operators forgot to indicate portability in these cases.

Table 2.1 below displays estimated portable machine counts within the NCCID training data, excluding historic images. For positive patients, there were *78* studies labelled with some reference to *portable* in their *Body Part Examined* DICOM attribute (*original counts*), accounting for approximately *1*.*4%* of X-ray studies. In comparison, the number of portable machines indicated by this DICOM attribute accounted for *0*.*9%* of negative patient studies. After extrapolating the *portable* status to all studies taken on the models where portability was indicated at least once, the proportion of X-ray studies taken on portable devices increased to approximately *14*.*3%* for positive patients and *16*.*7%* for negatives (*adjusted counts*).

**Table 2.1.** Estimated number of X-ray studies originating from either stationary or portable machines for COVID positive and negative patients.

| scanner type | COVID-positive |  | COVID-negative |  |
| --- | --- | --- | --- | --- |
|  | original count | adjusted count | original count | adjusted count |
| stationary | 5489 (98.6%) | 4770 (85.7%) | 5490 (99.1%) | 4610 (83.3%) |
| portable | 78 (1.4%) | 795 (14.3%) | 49 (0.9%) | 927 (16.7%) |

### 3. Cohort analysis

This section explores the geographic, demographic and temporal coverage of the NCCID database. The aim is to measure if/how the NCCID differs from the general COVID-affected population and how any disparities might limit the generalisability of AI solutions.

#### Geographic Coverage

Figure 3.1 details the number of patients submitted to the NCCID from each NHS England region (NHS, Regional teams n.d.) and Wales, split by their confirmed COVID-19 status, as measured via a RT-PCR swab test (positive = orange, negative = blue). The regional data was aggregated from the 19 sites that had submitted data by the analysis cut-off date.

**Figure 3.1.**
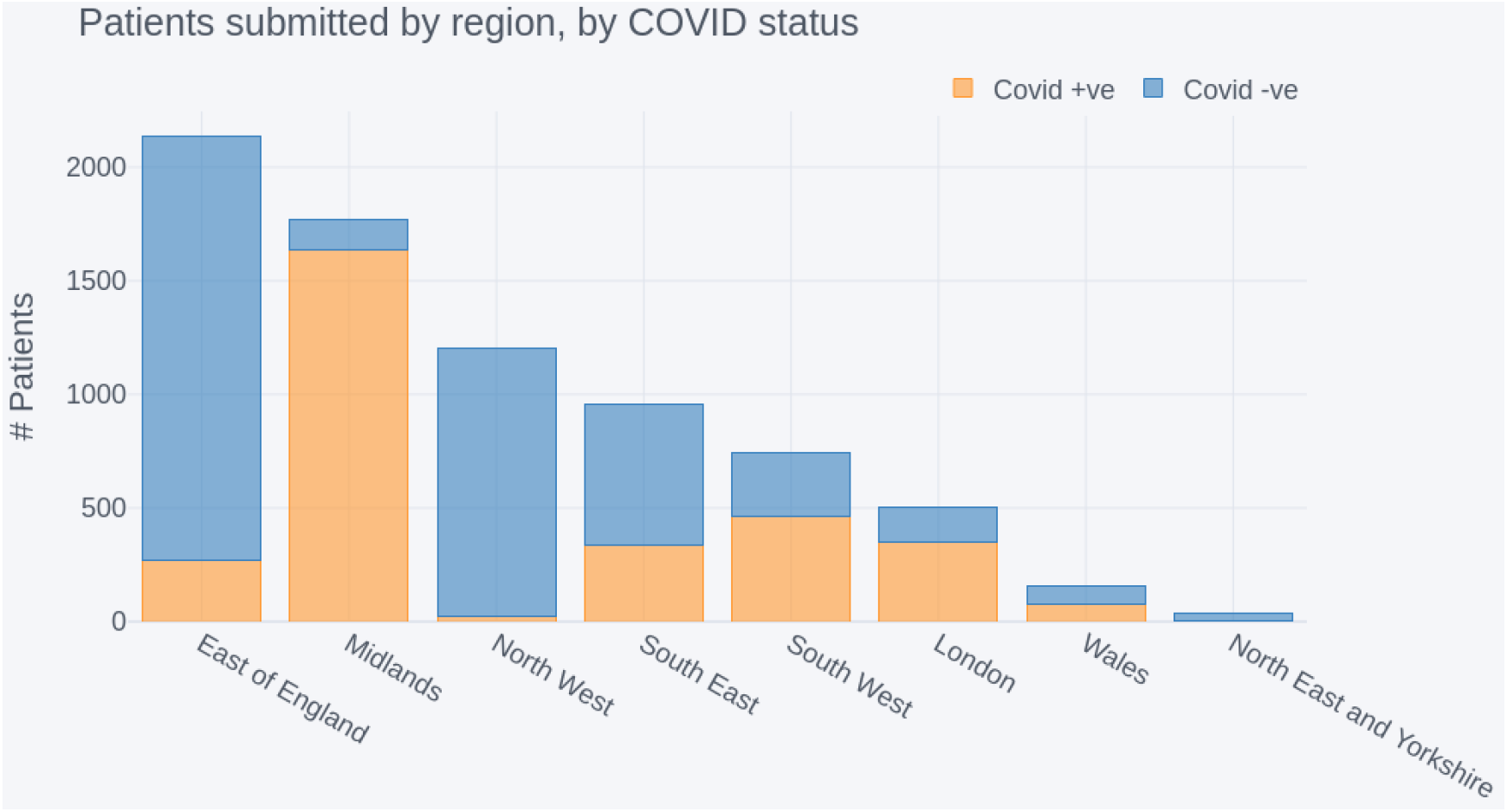
NCCID positive and negative patients submitted by region, sorted by total contribution.

In addition, Figure 3.2 displays two choropleth maps showing (A) the proportion of COVID-19 hospital admissions, within each NHS England region and Wales, as reported by Public Health England (PHE, coronavirus dashboard n.d.) and (B) the proportion of COVID-19 positive patients in the NCCID for the same geographic boundaries. Boundary data was sourced from the ONS geoportal (ONS, geography portal n.d.).

**Figure 3.2.**
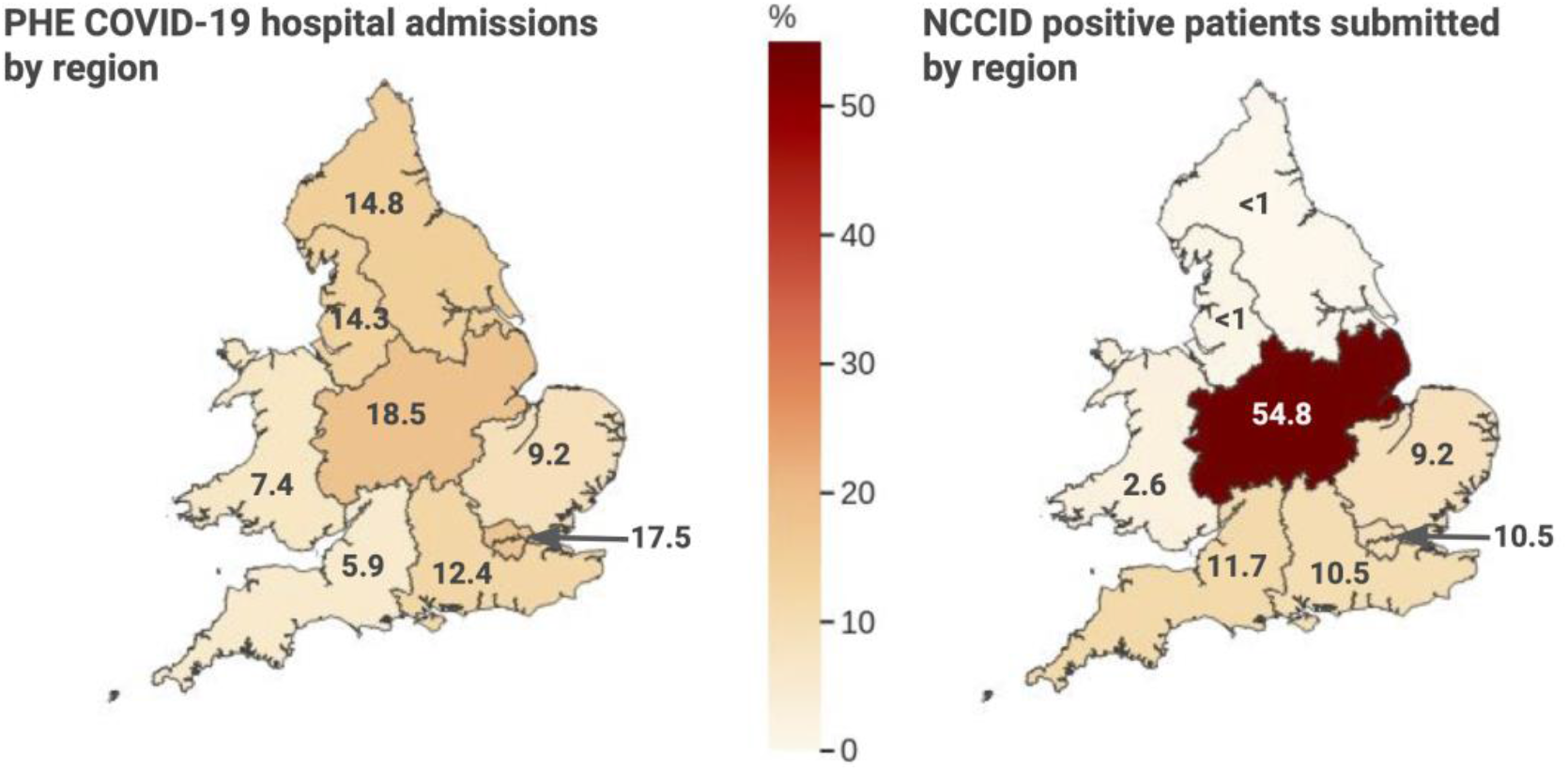
Comparison of national COVID-19 admissions at a regional level with NCCID positive cases

The highest proportion of data originated from the East of England region, which accounted for *2,134* patients in total. However, the vast majority of these (*1,862*) were negative patients, submitted by a single site. The second highest reporting region was the Midlands, with a combined total of *1,769* patients in the database. In contrast to the East of England, the vast majority of patients submitted in the Midlands were positive cases (*1,638*), and *1,511* of these originated from a single site.

Other regions submitted less data overall, but regions in the South of England (including London) and Wales had comparatively even contributions of positive and negative cases. Coverage of positive cases in the North of England and Yorkshire was limited, with the North East and Yorkshire region having only *33* patients in total.

The NCCID’s geographic coverage of COVID-19 patients was largely concentrated in the Midlands, accounting for *54*.*8%* of positive patients in the training data. After the Midlands, the East of England, London, South East and South West of England accounted for *41*.*6%* of positive patients in total (*9*.*2%, 10*.*2%, 10*.*5%*, and *11*.*7%*, respectively). Data from Wales, the North West, and the North East and Yorkshire regions collectively made up just *3*.*6*% of NCCID positive patients.

This was at odds with COVID-19 hospital admissions (as reported by PHE) which were more evenly spread across England and Wales. Specifically, London, the Midlands, North East and Yorkshire and the North West accounted for approximately *15-18%* of admissions each. Wales, the South East, East of England and South West accounted for smaller proportions of *10*.*3%, 9*.*8%, 7*.*0%* and *5*.*1%* of admissions, respectively.

#### Demographic Coverage

The purpose of this section is to establish how generally representative the NCCID cohort is of the population hospitalised due to COVID-19 and whether good representation carries through to the most severe outcomes (through the mortality variable). Understanding the underlying causes of any demographic differences in COVID-19 prevalence or outcomes is beyond the scope of this paper.

Subsequent to applying the cleaning and merging pipeline (see *clinical data* methods section), demographic data was available for *sex=85%, ethnicity=69%*, and *age=86%* of patients in the NCCID (*n=3,168*). Distributions of these categories within the NCCID were compared against reference datasets, where available, or COVID-related statistics reported by the International Severe Acute Respiratory and Emerging Infection Consortium (ISARIC) and the general UK population reported by the 2011 national census. Equivalent comparative data was not publicly available for Wales, as such, data from Welsh health boards is excluded from the subsequent demographic results. Comparisons were made for both admissions and mortality rates where the total sample size of patients with recorded deaths was *n=694*. In all subsequent comparison plots the NCCID is indicated using blue and comparative datasets are displayed in orange and green.

The NCCID is a subsample of the population that is hospitalised due to COVID-19, and a dynamic resource that will continue to grow over the coming months. It is sensible to assume that the sample of NCCID data being scrutinised in this paper will deviate from the final population of both the NCCID and general COVID-effected population. To account for some of this sampling error in the below comparisons, we applied a bootstrap method to generate confidence intervals for the NCCID data. The plotted proportions of a given category, e.g., percentage of patients aged 18-64, represent the median percentage across 1000 bootstrap samples. Similarly, error bars on the subsequent plots represent the 95% confidence interval (ci) of measurements across the bootstrap samples. In each case, the sample size of the bootstrapped distributions was equal to the size of the relevant original NCCID sample (i.e., if the original NCCID sample had n=3000 patients with sex data available then the bootstrapped samples each contained n=3000 entries).

##### Sex

Figure 3.3(A) compares the split of male (*n=1,797*) and female (*n=1,295*) positive cases within the NCCID to that of the general UK population via the 2011 national census (ONS, Census 2011 n.d.) *n=63,182,000*, and the COVID-effected population reported by ISARIC (Docherty 2020), *n=20,113*. At *58%* male to *42%* female (*ci = 56-60%male:40-44%female*), the NCCID was more closely aligned to the *60%:40%* ratio reported in COVID-19 admissions than the *51%:49%* split of the general UK population.

**Figure 3.3.**
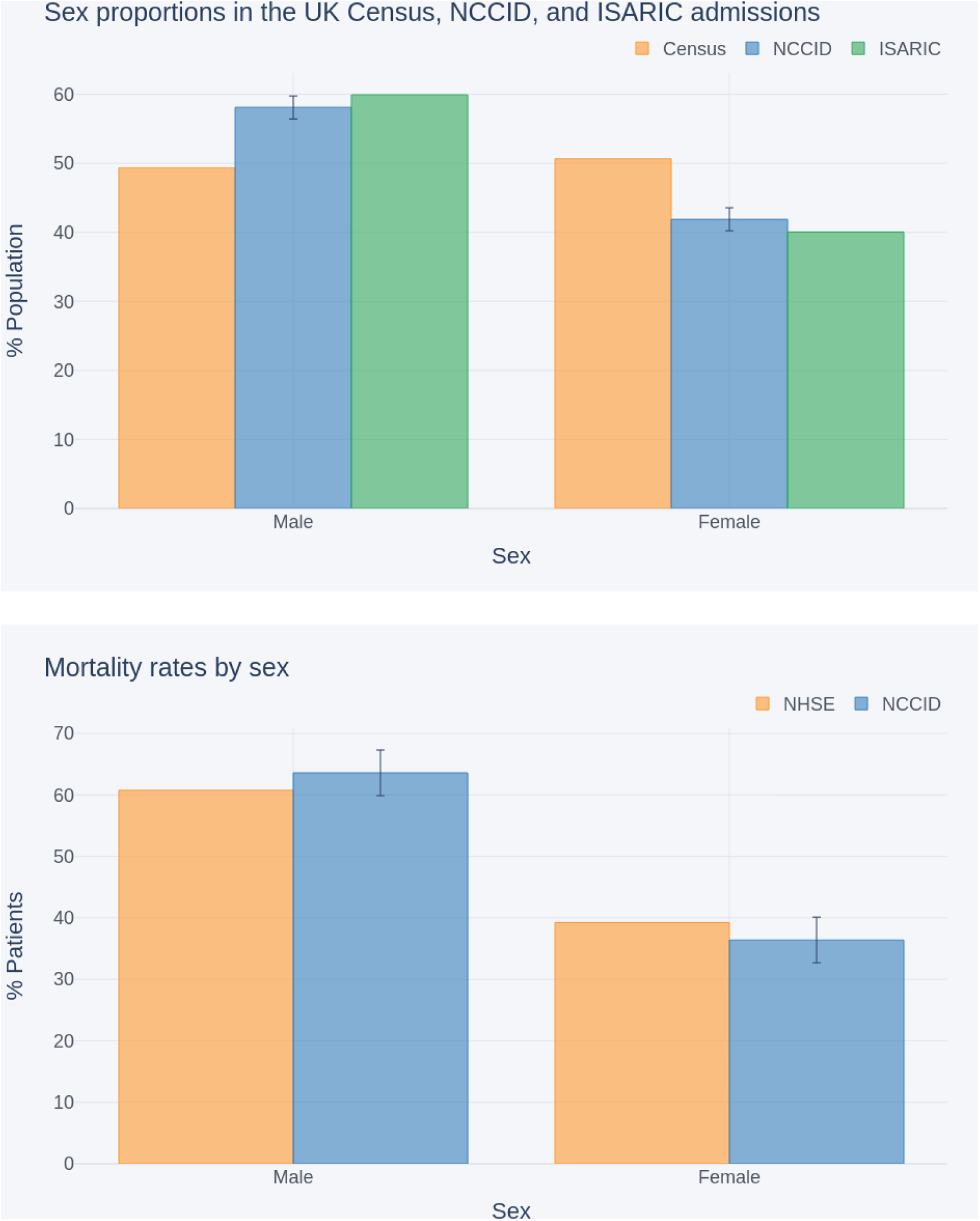
Comparison of sex split within: (A) the NCCID COVID-19 patients, the general UK population (as reported in the 2011 census) and COVID-19 hospital admissions (reported by ISARIC); (B) NCCID recorded deaths and NHS England COVID-19 hospital mortality data.

Figure 3.3(B) compares the male:female mortality rates within the NCCID cohort (*n=673*) against those reported by NHSE (*n=32,483*), up to the cut-off date, 29/10/2020 (NHS, COVID-19 Daily Deaths n.d.). The NHSE mortality data exhibited a male to female ratio of *61%:39%*. This fell within the 95% confidence interval for the NCCID, *60-67%:33-40%*.

##### Ethnicity

Figure 3.4(A) compares the ethnicity proportions (*Asian*, B*lack, Other, White*) of NCCID patients, *n=2854*, against the general UK population as reported in the 2011 UK census, *n=63,182,000*, (ONS, Census 2011 n.d.) and the COVID-affected population reported by ISARIC, *n=30,693* (Harrison 2020).

**Figure 3.4.**
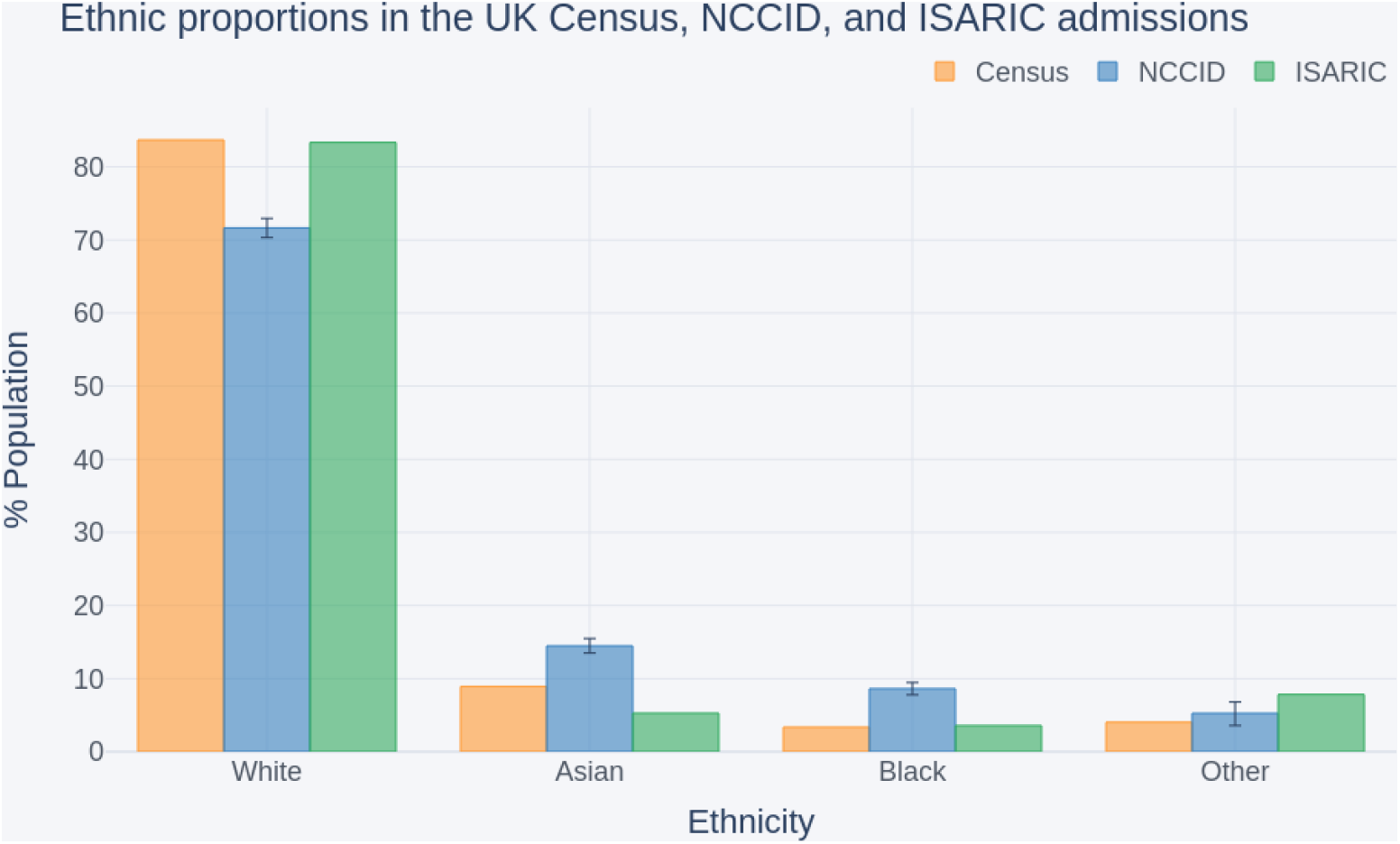
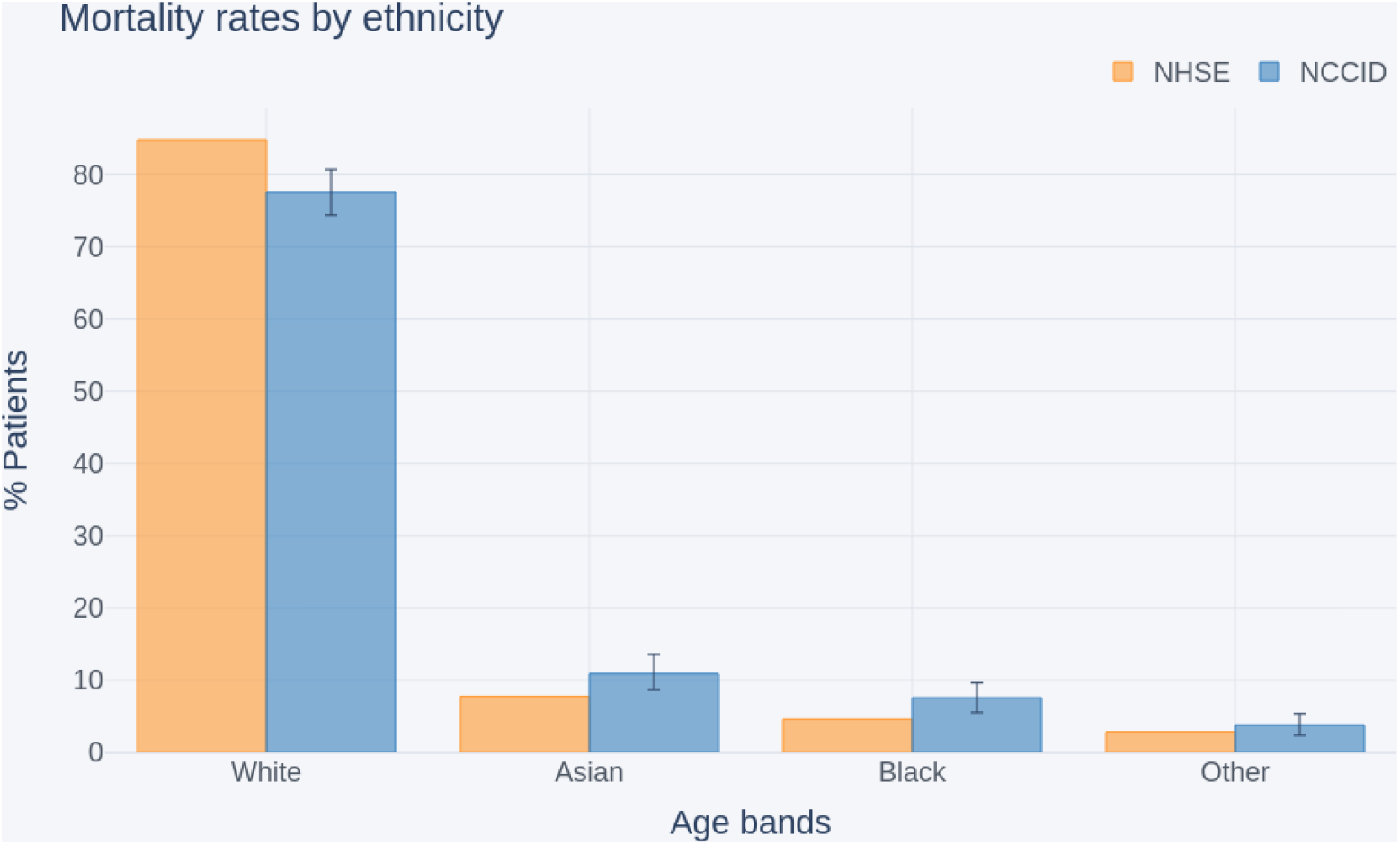
Comparison of ethnicity proportions within (A) the NCCID COVID-19 patients, the UK population (as reported in the 2011 national census) and COVID-19 hospital admissions (reported by ISARIC); (B) the NCCID recorded deaths and NHS England COVID-19 hospital mortality data.

The *White* group accounted for *83%* of individuals in both the census and ISARIC populations. In contrast, only *72%* (*ci = 70-73%*) of NCCID COVID-positive patients were from *White* ethnic backgrounds. This was counterbalanced by higher proportions of *Asian* (*median=14%, ci=13-16%*) and *Black* (*median=9%, ci=8-10%*) people, than observed in either the Census (*Asian = 9%, Black = 3%*) or ISARIC (*Asian = 5%, Black = 4%*). In addition, ISARIC reported higher proportions of patients from *Other* minority backgrounds (*8%*) than in NCCID (*median=5%, ci=4-6%*), whilst the census data indicated that approximately *4%* of the UK population belonged to this group.

Figure 3.4(B) compares the ethnicity proportions within the subset of NCCID patients that have recorded deaths and ethnicity data (*n=633*) to the ethnicity proportions reported by NHSE for COVID-19 in-hospital deaths in England (NHS, COVID-19 Daily Deaths n.d.), up to the reporting cut-off date (*n=29,610*).

Similar to the admissions data above, the NCCID mortality data was under-representative of the *White* ethnic group *(median=78% ci=74-81%*), and over-representative of the *Asian* (*median=11%, ci=9-13%*) and *Black* (*median=8%, ci=6-10%*) groups, compared to mortality rates in the broader COVID-population (*White=85%, Asian=8%, Black=5%*).

##### Age

Figure 3.5 compares the percentage of NCCID patients within a set of age bands (*0-5, 6-17, 18-64, 65-85, 85+*) to the percentages for COVID-19 hospital admissions across England, as reported by Public Health England (PHE, coronavirus dashboard n.d.). The comparisons are shown at both the national level as well as within each NHS England region.

**Figure 3.5.**
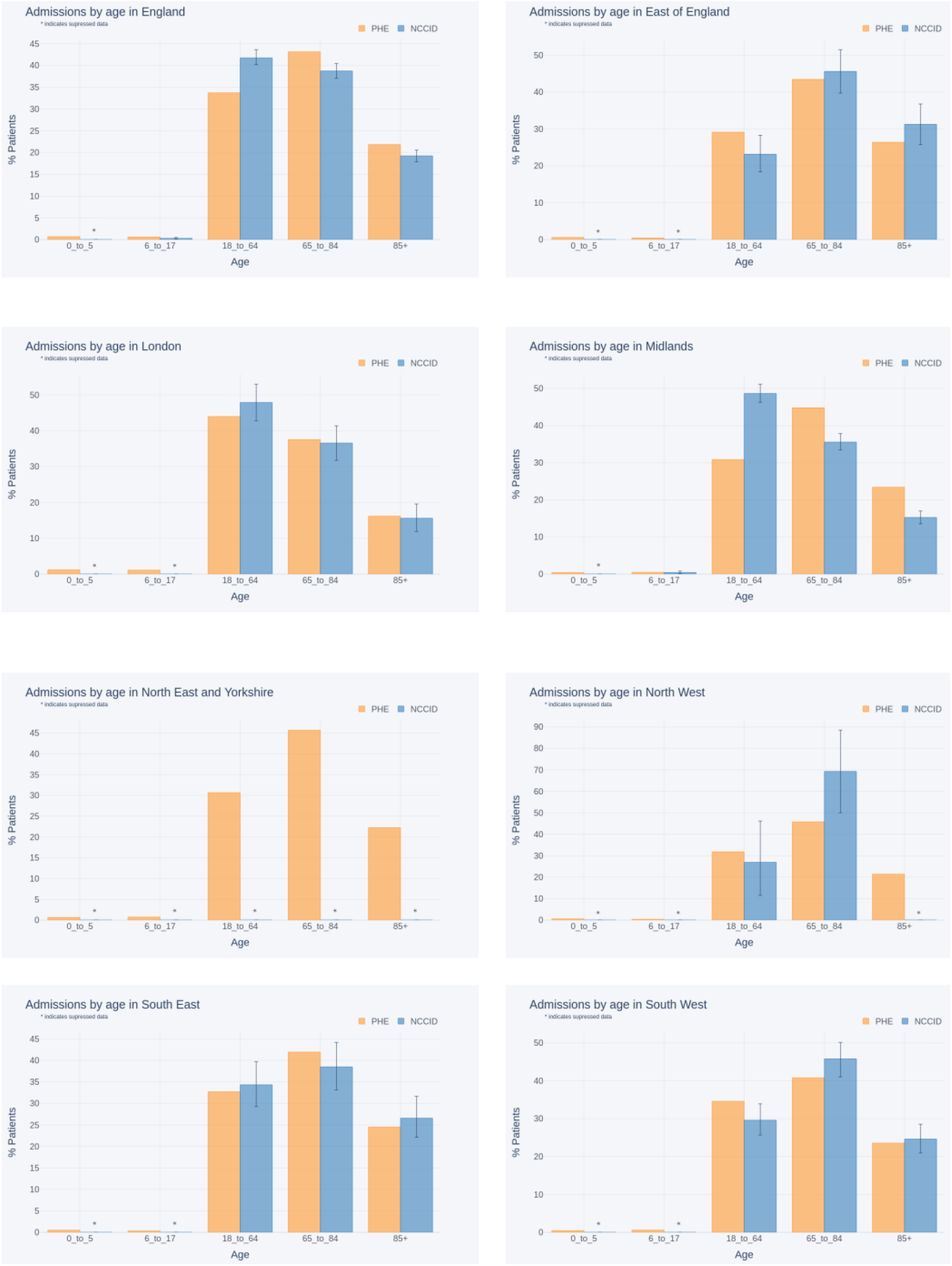
Comparison of age proportions between COVID-19 hospital admissions (reported by PHE) and NCCID positive patients for (A) England, (B) East of England, (C) London, (D) Midlands, (E) North East and Yorkshire, (F) North West (G) South East and (H) South West

As reflected in the geographic analysis, regions in the North of England had insufficient data to make meaningful comparisons. Specifically, data availability was below the suppression threshold in all age groups for the North East and Yorkshire and most age groups for the North West. The error bars for the remaining age groups in the North West, *18-64*, and *65-85*, spanned *30-34* percentage points respectively.

Amongst the regions that had enough data to support comparisons, most showed no statistically significant differences between the NCCID and PHE. For London (*n*_*PHE*_ = *25,804, n*_*NCCID*_ *= 353*) and the South East (*n*_*PHE*_ *= 15,690, n*_*NCCID*_ *= 335*) PHE data fell within the NCCID confidence intervals for all age-groups. The two data sets were closely aligned in the South West (*n*_*PHE*_*= 26,876, n*_*NCCID*_*= 463*), where only the *18-64* and *65-85* age bands fell outside the confidence interval by just *1%* each. Similarly, in the East of England (*n*_*PHE*_*= 11,252, n*_*NCCID*_*= 272*), the PHE data for the 1*8-64* age group was again just *1%* outside the upper bound for the NCCID, and all other age bands fell within the confidence interval.

The single exception was the Midlands, which exhibited a large difference of *18%* (*ci=15-20%*) between PHE (*n=26,661*) records and the NCCID (*n=1638*) for the *18-64* age band. This was counterbalanced by smaller proportions of over 65s than observed by PHE. These deviations can be reasonably attributed to the fact that data was collected by a single site, located in an urban area. Furthermore, given that the Midlands contributed a substantial volume of positive patients to the NCCID, this overrepresentation of *18-64* year olds extended to the national level comparison (*median*_*NCCID*_*= 42%, ci = 40-43%, n*_*NCCID*_*= 3088, median*_*PHE*_*= 33*.*7%, n*_*PHE*_*= 137,757*).

The NCCID had low numbers of patients in the 0-5 group at a national level, and low numbers for the 6-17 group in all geographies.

Figure 3.6 compares age breakdown of NCCID patients with recorded deaths to age breakdowns of in-hospital COVID-related deaths reported by NHSE (NHS, COVID-19 Daily Deaths n.d.). A different set of age bands were used to align to the NHSE data: *0 - 19, 20 - 39, 40 - 59, 60-79, 80+*.

**Figure 3.6.**
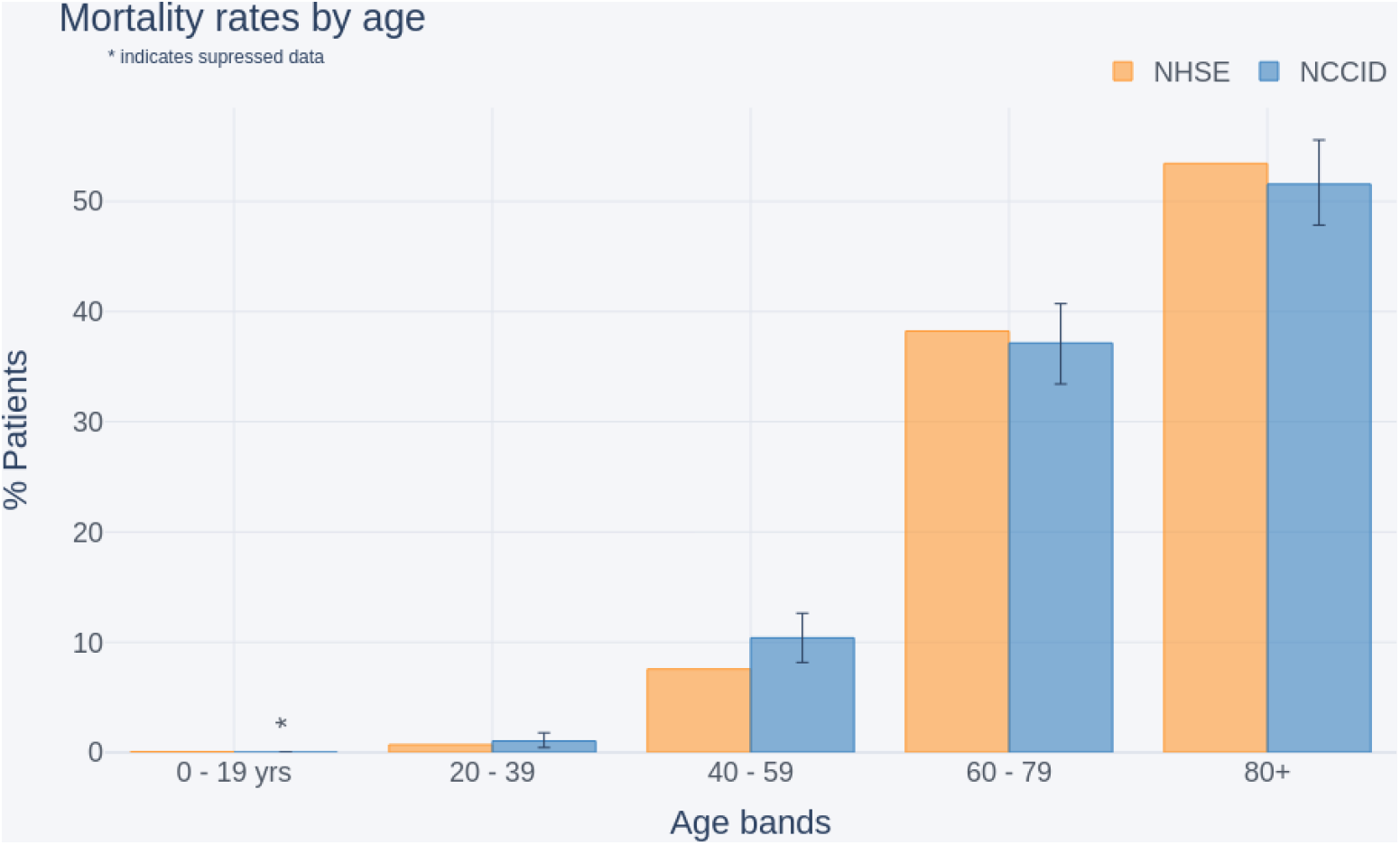
Comparison of age distributions between recorded COVID-19 deaths (as reported by NHSE) and the NCCID (England only).

Although the age bands used by NHSE *(n=32,484*) are different to those used in the admissions comparisons above, we can see a general knock-on effect, where over-representation of younger people in the dataset resulted in a larger percentage of *40-59* year olds with recorded deaths in the NCCID (median=10%, *ci=8-13%, NHSE=7%*).

#### Temporal coverage

This section investigates the approximate hospital admission dates of the NCCID patients to identify how well the NCCID has captured patients across the course of the pandemic. The total number of NCCID patients with a positive RT-PCR swab test occurring each week since 1 March 2020 was compared to the total number of confirmed COVID-19 patients admitted to hospital each week for the same period according to PHE data (PHE, coronavirus dashboard n.d.). This analysis was performed at a national level, including data across the whole of England and Wales. Given that there were (at the time of study) no NCCID sites in Scotland and Northern Ireland, data from these nations was omitted from PHE admissions calculations. The two time series are displayed in Figure 3.7.

**Figure 3.7.**
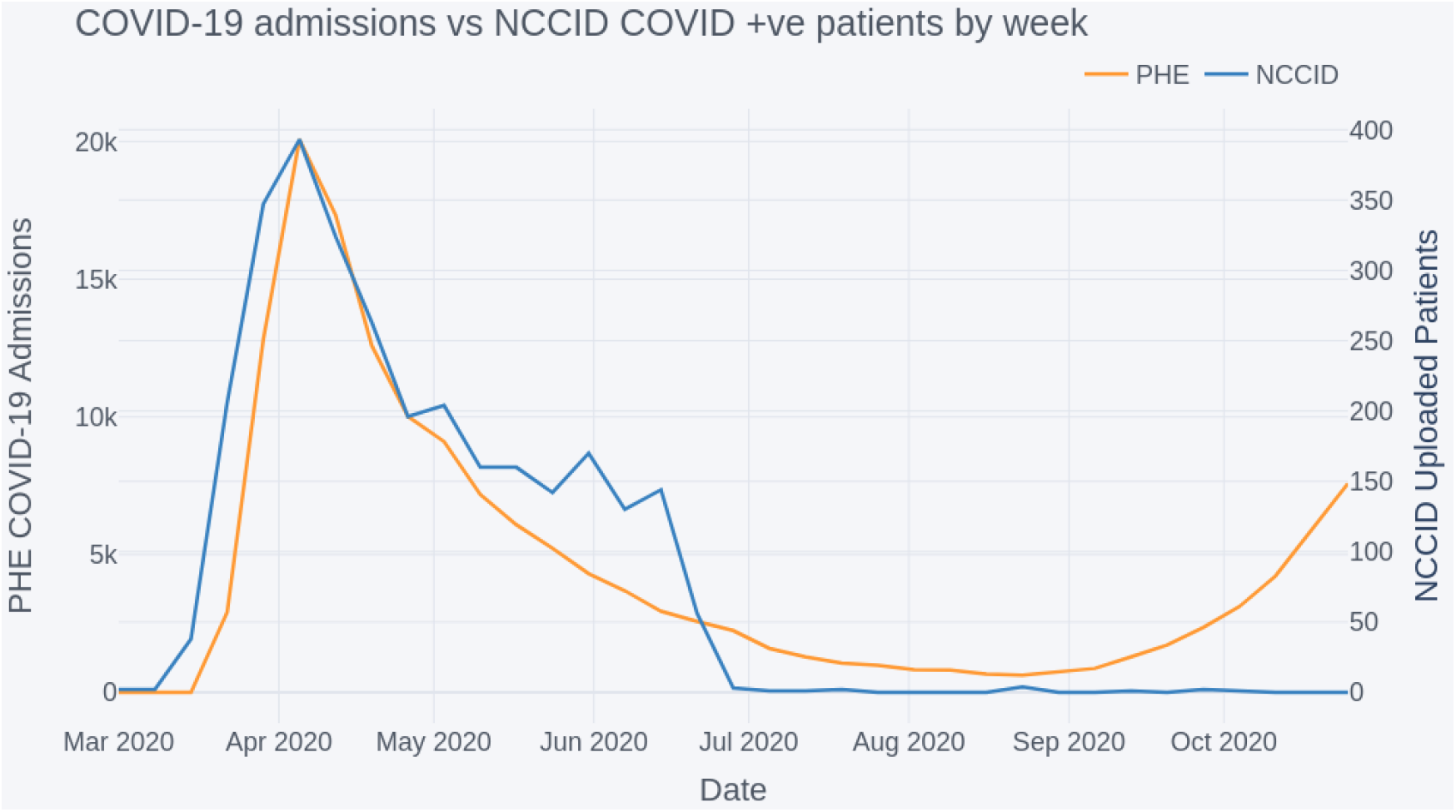
Comparison of COVID-19 admissions to NCCID positive cases by week

The peak of both datasets was aligned, occurring on 5 April, with a gradual decrease in numbers until the summer period, July to September 2020. From September onwards the national COVID-19 admissions began to rise again, however this was not (up to the analysis cut-off 29/10/20) reflected by a rise in positive patients admitted into the NCCID database.

## Discussion

### Findings of data completeness analysis

Clinical information is an important complement to the chest images. Gaps in the clinical information can deprive researchers of contextual data on the patient’s health for inclusion in analyses and ML models. For instance, incompleteness of the *FiO2* data may hinder the development of mortality or deterioration risk scores that take this field into account. Analogously, since clinical information may be used to control for confounders, missing entries can reduce a researcher’s ability to draw firm conclusions from the data.

The overall availability of clinical data varies by each field in the dataset. Key dates including when the RT-PCR swab was taken and when a patient was admitted to hospital are well covered, and can provide useful insight into the timelines of image acquisition during the patient care pathway (e.g., Figures 2.1 and 2.2).

The occurrence of pre-existing conditions is also relatively well characterised, particularly for cardiovascular and kidney diseases. This information should allow data users to account for the effects of comorbidities in their analyses, which have been shown to play a significant role in disease outcomes for COVID-19 patients (Guan 2020, Wang 2020, de Lucena 2020, Petrilli 2020).

Information relating to the patients’ conditions upon hospital admission (e.g., blood pressure and white-cell count) were the least well reported, with a mean of 65% null values in this category compared to 49% for dates, 53% for medical history, and 56% for COVID-19 fields. Data users should also be aware that the reporting units for these metrics may vary between sites, making it difficult to disambiguate overlapping values, and causing artificially high variances for some metrics (see Table C.1 of Appendix). To remedy this, we plan to make site-specific unit information available to users once collated, even though it is unlikely that all participating sites will be able to provide such information. It should also be noted that some of the missing data originates from the fact that specific hospitals do not commonly measure all of the listed metrics. For example, several sites report that they do not routinely measure *Troponin T* on admission. Furthermore, some fields such as *O2 saturation* are obsolete and no longer requested in the data collection spreadsheet.

Overall, the causes of missing information in the NCCID are difficult to identify because of their number and diversity. It is nevertheless known that the following factors have contributed to incompleteness of clinical data across the different categories:

- Staff at data-collection sites may have been unable to fill in certain fields due to time pressure and the emergency situation.
- Depending on the site, data has been gathered by staff (research nurses, radiologists, etc.,) with access to different clinical information systems and records. Therefore, the person collecting and uploading data to the NCCID may have been unable to get hold of specific clinical information.
- Certain fields could only be present in a relevant subset of patients, and were otherwise left empty. For example, a few fields referred to secondary RT-PCR swab tests (*date of acquisition, date of result, result*) and secondary chest X-rays (*date, severity*), which were only required, and consequently filled in for some patients. Additionally, the reporting of *date of death*, and stage of *chronic kidney disease* were much higher when selecting the subset of patients for whom death or presence of kidney disease had been reported. Similar effects are likely to be the underlying cause of the relatively high occurrence of missing values in COVID-19 fields such as *ITU admission, intubation* and *severity of disease* (BSTI n.d.) *in secondary images*.
- Information such as medical history may not have been provided by the patient, for example because they were incapacitated.
- Data may not have been gathered as part of routine clinical practice, see the above remarks.

Plans are in place to establish a link between the NCCID and ISARIC-4C (ISARIC n.d.) that will automatically populate clinical information for patients included in both datasets. This link aims to improve the availability of clinical data in the NCCID whilst relieving the burden on clinical staff to provide additional information.

### Findings of image characteristics analysis

#### Historic vs acute images

The number of total, acute or historic image studies varied across COVID-positive patients. In general, patients were less likely to have historic CT data available (*median=0 studies*), compared to X-ray (*median=1 study*). This is likely driven by the general disparities in availability between the two modalities, given that X-rays are faster and cheaper to acquire, and are therefore more frequently used in the UK clinical setting. Investigators that wish to incorporate historical data as a means of accounting for pre-existing pathologies or understanding longitudinal risk factors should possibly focus on X-ray studies.

Both X-ray and CT had a median of 1 study per patient, but there were many more X-ray studies available overall (approximately 12,000 compared to 1,500). It is sensible that researchers building diagnostic tools should focus on X-ray data, as these are also likely to be most useful in the UK clinical setting. However, given that CTs are likely to be used in the more severe/difficult cases, those wishing to analyse disease severity/prognosis can utilise CT data. One advantage of the CT data is that it provides much richer imaging information, encoded into a 3D volume where different view planes and slices through the relevant anatomy can be probed. In comparison, X-ray image resolution tends to be higher but only a single projection is possible.

The total number of MRI studies is currently too low (17 studies) to be useful in the machine learning setting. This is likely to remain true even as the database grows, as low numbers are caused by the rarer adoption of MRI in the treatment of COVID-19 patients, which in turn, limits the clinical relevance of this modality.

#### Acquisition timing

Analysis of image timings with respect to patient PCR-RT swab dates and onset of symptom dates revealed that X-rays were predominantly used at the early stages of a patient’s care pathway. Interestingly we identified the median offset between swab date and X-ray was −1 day, which suggests that X-rays were commonly being used as diagnostic aids. This is likely a result of limited testing capacity during the earlier stages of the pandemic. In contrast, CT images were generally used later in the care pathway, with greater variance between patients on the specific timing of scans. These findings reflect BSTI clinical guidelines for the UK, which stipulated that CT should be used sparingly as a diagnostic tool, to preserve capacity for normal operation (BSTI n.d.).

Concentrating on the response to COVID-19 in the UK and the NCCID, data users may want to focus on building diagnostic tools using X-ray images, and could potentially use CT scans to study disease severity, progression and prognosis. It remains to be seen whether improved testing capacity or other factors will modify the timings for either modality in the later stages of the pandemic, and therefore change the technological needs of the response to COVID-19 in the UK.

#### Scanner types

X-ray and CT images present in the NCCID were captured on a range of systems from multiple manufacturers, providing variability in the type of images available. This was true for both positive and negative patients, although the ratio of positive to negative varied somewhat by manufacturer. Users of NCCID should take into account the relative frequencies of imaging across the different manufacturers (and models) to minimise unwanted bias. For instance, Siemens is the dominant manufacturer for CT, but large amounts of X-ray data was available for a number of providers, which could help produce generalisable models.

Due to limitations imposed by the pandemic, it was suspected that imaging data in the NCCID would originate from a combination of portable and stationary X-ray machines. Portable machines are easier to quickly sanitise between sessions and could more readily be moved to quarantine wards as part of hospital infection control measures, making it possible that there would be a higher prevalence of such machines in the patient cohort (Kooraki 2020). Exploration of the DICOM headers initially identified a small proportion of positive scans (*1*.*4%*) acquired on portable devices, with just over half of this this percentage negative scans (*0*.*9%*). This was then extended to all studies taken on the same scanner models, such that *14*.*3%* of positive X-rays and *16*.*7%* of negative X-rays were estimated to come from portable machines. These preliminary findings do not suggest a large imbalance in the ratio of portable and non-portable scanners between the positive and control cohorts. However, in lieu of a more definitive method for identifying portable machines from DICOM information we estimated prevalence based on notes in the *Body Part Examined* attribute. It is plausible that this method under-estimates the true number of portable scanners, as such, further investigation of this issue is recommended. Examining a sample of images from the various devices may provide a more robust measure of portability for data users but the above analysis serves to highlight this aspect of the NCCID data.

Awareness of potential model confounders is crucial to ensure efficacy of ML models, particularly with respect to how performance generalises beyond the training data. For instance, significant disparities in the prevalence of certain equipment types between the positive and control cohorts could produce an ML model that successfully differentiates the two groups. However, is it conceivable that the decision boundaries in such a model are based on attributes of the medical imaging machinery (e.g., resolution, projection etc.) rather than disease related attributes. Data users should take care to balance their training samples, ensuring a good variety of scanner types within both cohorts, to build models that generalise well to the variety of clinical imaging equipment used in the UK. Indeed, there are many additional confounders to be aware of including but not limited to:

- *Digital radiography* (DR) vs *computed radiography* (CR) which are different techniques for digitising the X-ray signal, either directly from the panel (DR) or by scanning cassette-based phosphor storage plates into digital format (CR) (Table B.3 in Appendix B).
- *Photometric interpretation*, which refers to the image contrast such that *MONOCHROME1* scans should be inverted to match *MONOCHROME2* scans or vice versa (Table B.4 in Appendix B).
- *View positions*, e.g., *Anterior-Posterior* (AP), *Posterior-Anterior* (PA), *Lateral* (LL), etc., (Table B.5 in Appendix B).

By collecting data from multiple Trusts and Health Boards across the UK, the NCCID strives to provide a training database that can cover many of these confounding factors, and improve the efficacy of any resulting machine learning models in the clinical setting.

### Findings of cohort analysis

#### Geographic coverage

At time of analysis, the NCCID was not evenly sampled across the participating regions. We observed that COVID-19 positive-patients in the database largely originated from the Midlands, and very few patients originated from Wales and Northern England (Figure 3.1).

Several factors may underpin these disparities, including: 1) the number of NCCID sites within each region 2) the size and population coverage at each hospital site; 3) the number of positive COVID-19 cases recorded at each site; 4) the duration of time the site has been contributing to the NCCID for; and 5) the availability of research coordinators and PACS teams to upload all cases. Reason 3, is unlikely to be the driving factor, as indicated by Figure 3.2 in which PHE reported a more equal distribution of COVID-19 hospital admissions.

Low submissions from the North of England reflect the relatively small number of participating NCCID sites in these regions. The fact that the uptake of the programme has been uneven across different regions can be attributed to factors such as the reach of our professional network, constrained availability of staff to support our database, and variable responsiveness of local sites to national initiatives.

Regional disparities in the number of positive and negative cases submitted are more likely to be driven by factor 5, the capacity of PACS teams. The guidance given to hospital sites was to submit all positive cases with images taken in the acute setting, and a smaller sample of negative cases with acute imaging (approximately 100 per week if available). Due to the request for accompanying clinical data in positive cases, it is much easier for sites to submit negative cases, for whom only the images and a small number of clinical data points are required.

#### Demographic coverage

The NCCID aims to be a UK-wide initiative assembling a database that is as representative as possible of the entire population. Nevertheless, the present geographical coverage of the NCCID is partially skewed, which, if additional data curation is not applied rigorously, may produce biases in ML models trained on this resource. For example, issues may occur because of the incorrect representation of specific demographic groups and clinical risk factors such as pre-existing conditions (Docherty 2020, Pollán 2020). Indeed, we observed some of these downstream effects in the population analysis, particularly in the regional proportions of age-groups within the NCCID, which deviated most significantly from PHE data in the Midlands and Northern England. These effects accumulated in a general over-representation of younger adult patients compared to more elderly patients in the NCCID for both admissions and mortality.

In addition, the NCCID contains very low numbers of patients in the *0-5* and *6-17* age groups, partly because of the active omission of under-11s due to small counts, where the underlying cause is the low prevalence of symptomatic COVID-19 in children (Ludvigsson 2020, Dong 2020). Reduced availability of data for under-18s limits the use of the NCCID to adult diagnostic/prognostic models for the time being. This may change as the database grows, particularly as the exclusion of data from under-11s will be stopped once sufficiently high numbers are available.

The ethnic composition of the NCCID deviated from the 2011 UK census data. Whilst establishing the causes of this discrepancy would require additional investigation, the over-representation of *Asian* and *Black* groups for the admission data may, to some extent, be due to differences in the incidence of COVID-19. As a matter of fact, several studies have indicated higher corrected hospitalisation odds ratios for minority ethnic groups compared to people of white backgrounds (Martin 2020, Sze 2020, Harrison 2020, Docherty 2020). The reliability of the comparison between the NCCID and the census, however, is diminished by the fact that the latter is a decade old, so that more recent estimates (including the imminent 2021 national census) could exhibit a significant demographic shift in the benchmark for the UK population as a whole.

The comparison with ISARIC data was crucial for understanding how representative the NCCID is of the COVID-19 patient population that it is sampled from. Again, the NCCID displayed higher percentages of *Asian* and *Black* patients and lower percentages of *White* patients than the hospital admissions data from ISARIC. A similar effect was seen in the comparison with mortality data from NHSE.

The reasons why the NCCID diverges from other datasets in relation to ethnicity are not fully understood. Nevertheless, we believe that the most likely issue is the uneven geographical representation of the NCCID. This would be consistent with the fact that the *Asian* and *Black* groups are overrepresented, and the *White* group is underrepresented in every comparison of the NCCID with other nationwide datasets (UK census, NHSE and ISARIC). It is clear from the literature that the distribution of ethnicities in COVID related hospital admissions varies considerably between different regions (PHE 2020, Pollán 2020). For example, Sapey et al. (Sapey 2020), which looked specifically at COVID positive hospital admissions from around Birmingham saw a much higher proportion (*18*.*5%*) of patients of South Asian ethnicity. Apea et al. (Apea 2021), which carried out a similar analysis looking at COVID positive hospital admissions from around East London, saw a much higher proportion of patients of both South Asian and Black ethnicity (31% and 20% respectively). In an analogous way, the fact that a large fraction of the data in the NCCID has been collected in an urban area of the Midlands may have increased the representation of *Asian* and *Black* groups, and reduced that of the *White* group.

The male to female ratio of NCCID patients was found to closely align with the *60%:40%* split reported for COVID-patients by ISARIC. This is a departure from the approximately *50%:50%* split expected in the general population, as measured by the 2011 census data (where sex ratios are less likely to significantly vary over time, making the age of the census less of a limiting factor), and reflects findings of other COVID-19 studies (Gebhard 2020, Klein 2020, Petrilli 2020). A similar increased hazard ratio was observed in the male to female mortality rates, where the NCCID was well aligned to NHSE in hospital deaths data. Data users should be aware that there is a class imbalance (as is common in clinical studies) but unlikely to be severe enough to prevent the training of models that will generalise.

Overall, data users should keep in mind that, owing to the variable incidence of COVID-19, the NCCID is expected to have slightly different demographic composition to the general population. Several studies have reported different COVID-19 prevalence rates between men and women, ethnic groups and age groups (PHE 2020, Docherty 2020, Petrilli 2020, Gebhard 2020, Klein 2020, Sapey 2020, Sze 2020, Harrison 2020). As more sites are on-boarded and the database grows, we expect the composition of the NCCID to more closely reflect the populations reported by e.g., PHE, ISARIC, and NHSE. For the meantime, data users should be aware of these differences, and how underrepresentation of certain groups might affect model performance for those individuals. Whilst the risk of model unfairness relating to demographic disparities is less obvious in medical imaging than for other ML applications (e.g., facial recognition for law enforcement (Fussey 2019)), it is probable that disease manifestation differs across age groups and ethnicities, or that the prevalence of comorbidities varies across ethnicities and between urban and non-urban populations. Therefore, these characteristics may still have negative effects on the fairness of ML models. Furthermore, disease-related class imbalances play a relevant role in quantifying algorithmic bias, where fairness definitions based on pure demographic parity (Begley 2020) (Mehrabi 2019) may provide misleading measures of success and failure in this problem space, unless corrected to the relevant ratios.

#### Temporal coverage

The low numbers of positive cases uploaded to the NCCID training dataset since September 2020 suggest that the data capture pipelines were (up to the analysis cut-off in October) still processing the large backlog of patients from the first wave of the pandemic. Users should note that ML models built from the training data will capture the characteristics of the first peak, and may not generalise completely to patients admitted during the subsequent winter peaks, particularly in view of the emergence of a new strain of SARS-CoV-2, lineage B.1.1.7 (Andrew Rambaut 2020). Failures to generalise over time could arise from several factors, including:

- potential changes to disease manifestation associated with the new strain of SARS-CoV-2 that has dominated prevalence in the UK starting from December 2020 (Kirby 2021, Volz 2021), though such effects are speculative at the time of publishing;
- the prevalence of flu-related comorbidities, expected to be more common in winter months;
- any changes in the use of imaging for diagnostic/prognostic purposes between the early stages and later stages of the pandemic;
- changes to treatment policies over time (such as the introduction of dexamethasone) and how these affect disease severity;
- the roll-out of the COVID-19 vaccination programme, which in the UK has begun on 8 December 2020 (BBC 2020), and has delivered almost 18 million first doses (GOV 2020) at the time of writing;
- changes to non-pharmaceutical interventions (behavioural restrictions like lockdowns) and the down-stream effects these have on which members of the population are exposed to the virus.

It is noteworthy that COVID-19 admissions for the general population peaked at approximately 20,000 per week (for the period and regions studied in this article), whilst the peak of positive patients in the NCCID was orders of magnitude lower, at just under 400. Any statistics or models derived from the NCCID database are therefore likely to suffer from sampling error, which should be considered when reporting such analyses.

### Next Steps

The NCCID has made significant progress within the space of a few months to collect a sizable dataset to support research into COVID-19. However, there are a number of next steps, summarised below, which the NCCID initiative aims to implement in the short-to-medium term in order to better support data users:

1. We will re-engage with existing hospital sites to understand the reasons behind a decline in submission of recent cases and implement mitigating actions (see point 5).
2. We will engage new sites across the UK, focusing on rural and other underrepresented geographies, such as the North of England, Wales, Northern Ireland (point 4) and Scotland (point 3) to expand the geographic and demographic coverage of the NCCID.
3. We will implement a linkage with the Scottish National PACS and Safe Haven Network and
4. In Northern Ireland we will start by establishing a linkage with the Northern Trust PACS team.
5. We will implement a connection with the ISARIC-4C (ISARIC n.d.) dataset to improve the completeness of the clinical data fields while reducing the burden on hospital staff, since the data is linked across as opposed to collected afresh. It is hoped that lighter data-gathering processes will attract new sites, and motivate existing ones to contribute even more to the database.
6. We will carry out investigative work beyond clinical variables and metadata into the quality of the images themselves so as to assess their utility for algorithmic development.
7. We will implement automation pilots in a selection of sites to establish a continuous feed of images for positive and negative patients. Clinical data for these sites will be provided through the ISARIC-4C linkage.

### Conclusion

This paper aimed to provide further detail on the content of the NCCID’s training dataset, in order to support existing data users with their research efforts, raise awareness for the NCCID as a valuable resource that others may want to access, and inform both existing and potential data users of improvements we aim to make in future. The decision to publish this paper now, rather than after the improvements have been made, reflects the iterative nature of this particular initiative, and the urgency presented by the pandemic to ensure information is made available as quickly, transparently and securely as possible. The NCCID initiative has collected a large volume of imaging and clinical data within a short period of time; this has been achieved through the expertise of NCCID partners, lean agile delivery methods, and the prioritisation of COVID-19 response work. However, there are a number of considerations in the NCCID training dataset to be aware of, namely: 1) the limitations of its geographic and, consequently, demographic representation; 2) issues with clinical data quality and completeness. We have identified a number of improvements to address these considerations, and will continue to expand and refine the quality of the NCCID training dataset as an important tool in supporting the global response to COVID-19.

## Data Availability

Access to the dataset can be sought via an application to the National COVID-19 Chest Imaging Database (NCCID) Data Access Committee as described on the NCCID website linked.

https://nhsx.github.io/covid-chest-imaging-database/

## Supplementary resources

Additional information on the NCCID, including an overview of participating sites, existing data processors, live updates on the size of the training data and instructions for requesting access are all available through the main webpage.

More information on guidelines and schemas for the clinical data are available [through RSNFT](https://medphys.royalsurrey.nhs.uk/nccid/guidance.php), further detail is also provided through the [HDRUK portal](https://web.www.healthdatagateway.org/dataset/31f0148b-f965-4136-ab39-6c5bbbf8c2d9). The open-source data cleaning pipeline can be found on [NHSX github](https://github.com/nhsx/nccid-cleaning).

The codebase for the data warehouse is also open source and available through the [NHSX github](https://github.com/nhsx/covid-chest-imaging-database).

## Acknowledgements

The authors would like to thank the following individuals for their contributions to this work: Ayub Bhayat, Hena Aziz, Zain Eisa, Rob Howieson, Alison Lowe, Aliya Rafique, Anastasios Sarellas and Giuseppe Sollazzo. Joseph Jacob was supported by a Wellcome Trust Clinical Research Career Development Fellowship (209553/Z/17/Z) and by the NIHR BRC at UCL.

### Appendix

#### A. Glossary

##### Imaging study

a study is either an X-ray, CT or MRI examination. Studies may contain multiple images (series) if i.e., several projections, body positions, fields of view, contrast types etc., were acquired during the appointment, but all of these individual images (image volumes for 3D modalities) are all amalgamated to a single data point in time - a study. Each study is assigned a unique accession number on PACS.

##### Site Definition

when referring to a ‘site’ in this paper we are talking about a Submitting Centre for NCCID. This is a Trust in England, or a Health Board in Wales. A single Trust or a Health Board can contain many different contributing hospital sites.

Sites contributing as of 29th October 2020:

- [Ashford and St Peter's Hospitals NHS Foundation Trust](http://www.ashfordstpeters.nhs.uk/)
- [Betsi Cadwaladr University Health Board](https://bcuhb.nhs.wales/)
- [Brighton and Sussex University Hospitals NHS Trust](https://www.bsuh.nhs.uk/)
- [Cambridge University Hospitals NHS Foundation Trust](https://www.cuh.nhs.uk/)
- [Cwm Taf Morgannwg University Health Board](https://cwmtafmorgannwg.wales/)
- [George Eliot Hospital NHS Trust](http://www.geh.nhs.uk/)
- [Hampshire Hospitals NHS Foundation Trust](https://www.hampshirehospitals.nhs.uk/)
- [Imperial College Healthcare NHS Trust](https://www.imperial.nhs.uk/)
- [Liverpool Heart and Chest Hospital NHS Foundation Trust](https://www.lhch.nhs.uk/)
- [London North West University Healthcare NHS Trust](https://www.lnwh.nhs.uk/)
- [Norfolk and Norwich University Hospitals NHS Foundation Trust](http://www.nnuh.nhs.uk/)
- [Oxford University Hospitals NHS Foundation Trust](https://www.ouh.nhs.uk/)
- [Royal Cornwall Hospitals NHS Trust](https://www.royalcornwall.nhs.uk/)
- [Royal Surrey NHS Foundation Trust](https://www.royalsurrey.nhs.uk/)
- [Royal United Hospitals Bath NHS Foundation Trust](https://www.ruh.nhs.uk/)
- [Sandwell and West Birmingham NHS Trust](https://www.swbh.nhs.uk/)
- [Sheffield Children's NHS Foundation Trust](https://www.sheffieldchildrens.nhs.uk/)
- [Somerset NHS Foundation Trust](https://www.somersetft.nhs.uk/)
- [West Suffolk NHS Foundation Trust](https://www.wsh.nhs.uk/Home.aspx)

The NCCID are continually onboarding new sites, a up to date list of submitters is available on the NCCID information page^1^

#### B. Scanner and image types

##### Models

The full list of model types for X-ray and CT across all imaging available in the dataset are included in Tables B.1 and B.2 in the Appendix where counts below 100, have been suppressed for brevity but all the model names have been listed below.

A single model type accounts for all X-ray studies from the largest supplier: *Siemens - fluorospot compact FD*. For the second largest X-ray supplier, *Fujifilm*, the model attribute was universally missing, making it hard to determine if single or multiple models are present. Most other X-ray manufacturers had multiple models available in the dataset. In the case of CT studies, the dominant manufacturer (*Siemens*) was split across ten different models, two of which had more than 500 studies each.

**Table B.1.** Model Manufacturer Name (X-ray). Machines that are suspected to be portable have been highlighted in bold red.

| Manufacturer - Model (X-ray) | Studies |
| --- | --- |
| siemens - fluorospot compact fd | 5093 |
| fujifilm corporation - missing | 3560 |
| philips medical systems - digitaldiagnost | 2414 |
| **carestream health - drx-revolution** | 1050 |
| agfa - 3543eze | 719 |
| **philips medical systems - mobilediagnost wdr** | 538 |
| ge healthcare - optima xr220 | 526 |
| ge healthcare - discovery xr656 hd | 277 |

<sup>1</sup> https://nhsx.github.io/covid-chest-imaging-database/index.html

|  |  |
| --- | --- |
| philips medical systems - digitaldiagnost c90 | 249 |
| **kodak - cr850a** | 224 |
| samsung electronics - gm85 | 211 |
| **carestream health - cr975/cr0975** | 188 |
| carestream health - drx-evolution | 179 |
| samsung electronics - gr40cw | 174 |
| ge healthcare - definium 6000 | 168 |
| varian - varian_4336 | 151 |
| agfa - cr 35 | 143 |
| kodak - dr 7500 | 132 |
| agfa - cr30-x | 117 |
| agfa - cr 85 | 115 |
| ge healthcare - thunder platform | 110 |
| varian - varian_4343r | 110 |

For counts of studies below 100, we also have X-ray examples from:
**agfa** - cr15-x, dx-g, dxd30\_wireless, dxd40\_1000c, pixium\_4343e\_csi
**canon inc.** - axiom-multix m, cxdi, cxdi control software ne
**carestream health** - classic cr, **cr0850a**, drx-1, elite cr
**drgem** - gxr system
**ge healthcare** - discovery xr656, geode platform, global 1 platform, **wdr1**
**kodak** - classic cr, **cr850**, cr950, **cr975/cr0975**, drx-evolution, elite cr
**konica minolta** - 0862, cs-7
**philips medical systems** - pcr eleva

**Table B.2.** Model Manufacturer Name (CT)

| Manufacturer - Model (CT) | Studies |
| --- | --- |
| siemens - somatom definition as | 624 |
| siemens - somatom definition flash | 568 |
| siemens - somatom definition as+ | 412 |
| siemens - somatom drive | 200 |
| siemens - somatom definition edge | 141 |
| siemens - somatom perspective | 109 |
| siemens - somatom force | 103 |

For counts of studies below 100, we also have CT examples from:
**canon medical systems** - aquilion one, aquilion prime sp
**ge medical systems** - discovery 710, discovery ct750 hd, lightspeed16, lightspeed vct, optima ct660, revolution ct, revolution evo
**philips medical systems** - brilliance 64, ict 256, ingenuity core 128, ingenuity ct
**siemens** - emotion 16 (2007), sensation 64, somatom edge plus, somatom go.top, symbia intevo 16
**toshiba** - aquilion, aquilion one, aquilion prime, aquilion prime sp

##### X-ray format

**Table B.3.** The number of X-ray studies in Digital Radiography (DX) and Computed Radiography (CR) formats as identified from the Modality DICOM attribute (0008, 0060) for COVID positive and negative patients.

| Modality (X-ray Studies) | Positive | Negative |
| --- | --- | --- |
| DX (Digital Radiography) | 5026 | 4056 |
| CR (Computed Radiography) | 6699 | 1476 |

##### Photometric interpretation

**Table B.4.** The number of MONOCHROME1 and MONOCHROME2 X-ray studies as identified by the Photometric Interpretation attribute (0028, 0004) for COVID positive and negative patients. Images identified as MONOCHROME1 will need to be inverted to be aligned with those with MONOCHROME2.

| Photometric Interpretation (X-ray Studies) | Positive | Negative |
| --- | --- | --- |
| MONOCHROME2 | 8254 | 2562 |
| MONOCHROME1 | 3478 | 2971 |

##### View position

**Table B.5.** The number of X-ray studies by View Position attribute (0018, 5101) for COVID positive and negative patients e.g., Anterior-Posterior (AP), Posterior-Anterior (PA), Lateral (LL).

| View Position (X-ray Studies) | Positive | Negative |
| --- | --- | --- |
| Missing | 6662 | 2692 |
| AP | 3076 | 1598 |
| PA | 2003 | 1394 |
| OTHER | 10 | < 7 |

#### C. Clinical Data

**Table C.1.**
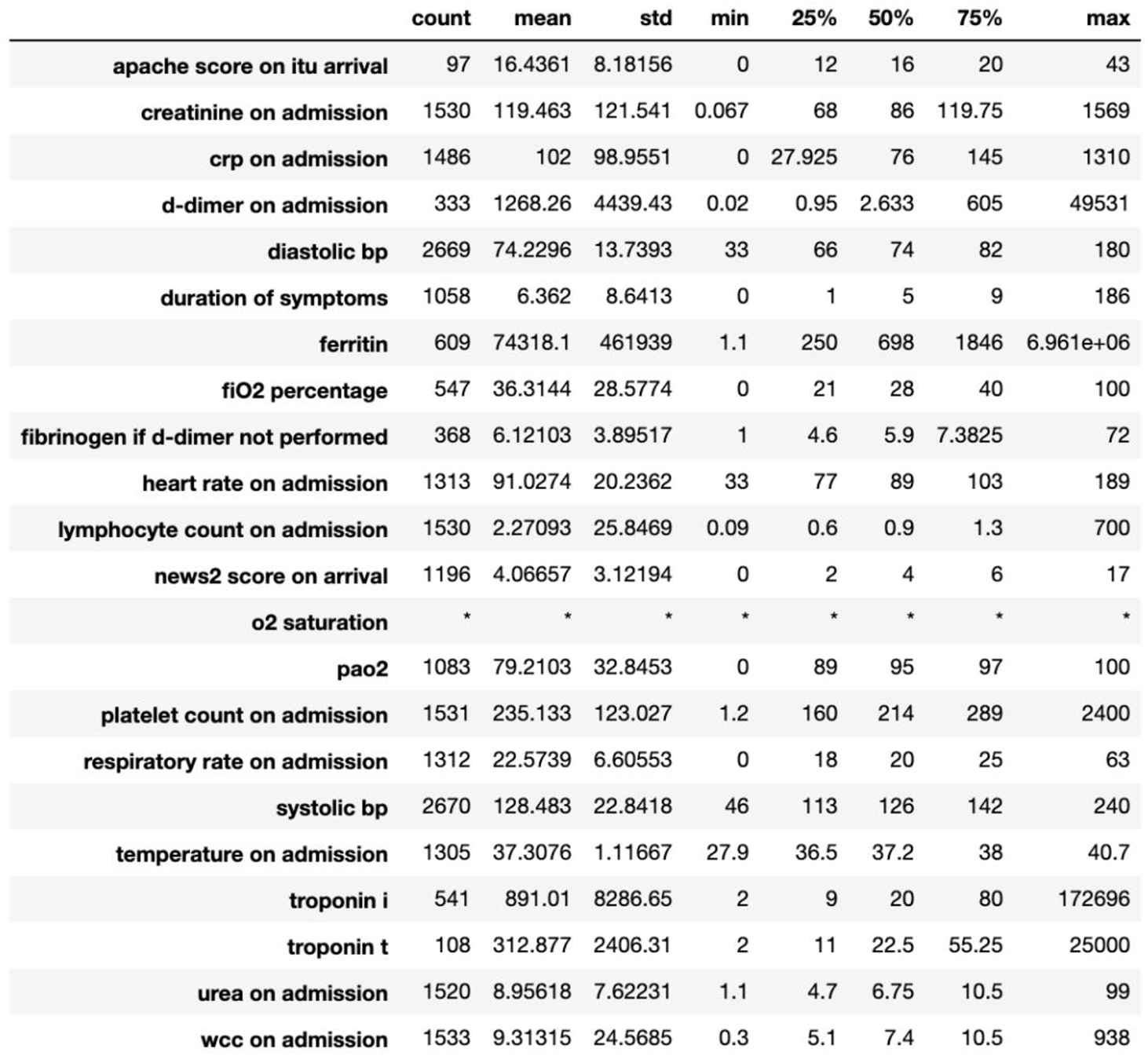
shows the summary statistics for the clinical admission metric (as of 29 October 2020). suppression of low counts is indicated by an asterisk (*).

|  | count | mean | std | min | 25% | 50% | 75% | max |
| --- | --- | --- | --- | --- | --- | --- | --- | --- |
| apache score on itu arrival | 97 | 16.4361 | 8.18156 | 0 | 12 | 16 | 20 | 43 |
| creatinine on admission | 1530 | 119.463 | 121.541 | 0.067 | 68 | 86 | 119.75 | 1569 |
| crp on admission | 1486 | 102 | 98.9551 | 0 | 27.925 | 76 | 145 | 1310 |
| d-dimer on admission | 333 | 1268.26 | 4439.43 | 0.02 | 0.95 | 2.633 | 605 | 49531 |
| diastolic bp | 2669 | 74.2296 | 13.7393 | 33 | 66 | 74 | 82 | 180 |
| duration of symptoms | 1058 | 6.362 | 8.6413 | 0 | 1 | 5 | 9 | 186 |
| ferritin | 609 | 74318.1 | 461939 | 1.1 | 250 | 698 | 1846 | 6.961e+06 |
| fiO2 percentage | 547 | 36.3144 | 28.5774 | 0 | 21 | 28 | 40 | 100 |
| fibrinogen if d-dimer not performed | 368 | 6.12103 | 3.89517 | 1 | 4.6 | 5.9 | 7.3825 | 72 |
| heart rate on admission | 1313 | 91.0274 | 20.2362 | 33 | 77 | 89 | 103 | 189 |
| lymphocyte count on admission | 1530 | 2.27093 | 25.8469 | 0.09 | 0.6 | 0.9 | 1.3 | 700 |
| news2 score on arrival | 1196 | 4.06657 | 3.12194 | 0 | 2 | 4 | 6 | 17 |
| o2 saturation | * | * | * | * | * | * | * | * |
| pao2 | 1083 | 79.2103 | 32.8453 | 0 | 89 | 95 | 97 | 100 |
| platelet count on admission | 1531 | 235.133 | 123.027 | 1.2 | 160 | 214 | 289 | 2400 |
| respiratory rate on admission | 1312 | 22.5739 | 6.60553 | 0 | 18 | 20 | 25 | 63 |
| systolic bp | 2670 | 128.483 | 22.8418 | 46 | 113 | 126 | 142 | 240 |
| temperature on admission | 1305 | 37.3076 | 1.11667 | 27.9 | 36.5 | 37.2 | 38 | 40.7 |
| troponin i | 541 | 891.01 | 8286.65 | 2 | 9 | 20 | 80 | 172696 |
| troponin t | 108 | 312.877 | 2406.31 | 2 | 11 | 22.5 | 55.25 | 25000 |
| urea on admission | 1520 | 8.95618 | 7.62231 | 1.1 | 4.7 | 6.75 | 10.5 | 99 |
| wcc on admission | 1533 | 9.31315 | 24.5685 | 0.3 | 5.1 | 7.4 | 10.5 | 938 |

## Footnotes

1 https://nhsx.github.io/covid-chest-imaging-database/index.html

